# Urinary vesicle biomarkers and kidney function – Results from the German AugUR study

**DOI:** 10.64898/2026.01.12.26343937

**Authors:** Luisa Schnobrich, Hannah C. de Hesselle, Lorena Mornhinweg, Rike Felgner, Martina E. Zimmermann, Caroline Brandl, Iris M. Heid, Hayo Castrop, Klaus J. Stark

## Abstract

**Background:** Progression into more severe stages of chronic kidney disease (CKD) based on estimated glomerular filtration rate (eGFR) and albuminuria are associated with increased risk of end-stage renal failure, cardiovascular diseases, and mortality. Vesicles in the urine are cell-derived particles containing constituents of the cells of origin. Little is known about the prognostic capacity of urinary vesicles for CKD progression.

**Purpose:** To evaluate the association between components of urinary vesicles and incident CKD.

**Methods:** In the AugUR study, a prospective population-based cohort study in individuals aged 70-95 years at baseline, we isolated and characterized urinary vesicles from 580 participants at two timepoints. In cross-sectional data, influences of age, sex and established kidney biomarkers on vesicular albumin and podocalyxin were characterised. Longitudinal data were used to test associations of vesicular albumin and podocalyxin with incident eGFR-based CKD and albuminuria.

**Results:** Cross-sectionally, urinary vesicle albumin and urinary vesicle-bound podocalyxin were moderately correlated with each other and with urinary albumin and alpha1-microglobulin, but not with eGFR. Vesicular albumin concentrations were influenced by sex, whereas age showed an effect on podocalyxin. After adjusting for age and sex, higher vesicular albumin was associated with higher urinary albumin and lower eGFR. Higher vesicular podocalyxin concentrations were associated with higher urinary albumin but not with eGFR. Both markers showed identical associations with urinary alpha1-microglobulin. In longitudinal analyses, baseline vesicular albumin showed association with incident CKD based on eGFR. This association was no longer present after adjustment for baseline eGFR. In contrast, higher baseline podocalyxin concentrations were predictive for decreased risk of incident albuminuria after adjustment for baseline free urinary albumin. Baseline-adjusted change in urinary vesicle albumin and urinary vesicle-bound podocalyxin were both associated with incident albuminuria, independent of free urinary albumin and other kidney biomarkers. Here, increase in follow-up versus baseline values were associated with higher risk for incident albuminuria, with higher effect sizes for vesicular albumin.

**Conclusion:** This study indicates that higher vesicular podocalyxin at baseline might be a potential predictor for lower risk for albuminuria over three years in an old-aged cohort. In contrast, longitudinal increase in urinary vesicle biomarkers, especially vesicular albumin, might be diagnostic markers for incident albuminuria in the elderly.

**KEY Messages:** What is known

- According to a previous study in animals, accumulation of albumin in the subpodocyte space leads to subsequent endocytosis by the podocytes.
- Podocyte-produced vesicles contain potential biomarkers of the deterioration of kidney function in humans.

What is new

- Biomarkers from urinary vesicles can be quantified from biobanked human samples.
- Higher vesicular podocalyxin at baseline might be a potential predictor for lower risk for albuminuria over three years in an old-aged cohort.
- Changes in urinary vesicle biomarkers over time, especially vesicular albumin, are associated with incident albuminuria independent of eGFR and free urinary albumin.

## INTRODUCTION

Chronic kidney disease (CKD), which is defined by reduced kidney function and/or kidney damage persisting for over three months [1], is a serious and growing global health problem, affecting more than 10% of the adult global population [2]. Kidney function is assessed by the determination of the glomerular filtration rate (GFR), estimated with serum biomarkers creatinine or cystatin C (eGFR) and by the degree of albuminuria, measured as the urinary albumin-creatinine-ratio (uACR) [1]. The urinary albumin excretion is determined by the degree of glomerular albumin filtration and the extent of tubular reabsorption, predominantly in the proximal tubule [3]. Based on its size and charge specificity, the intact glomerular filtration barrier (GFB) is largely impermeable for macromolecules, such as albumin. The GFB consists of three layers: the glomerular endothelial cells, basement membrane and podocytes. The latter are important for the function of the GFB, as they form a size barrier with slit diaphragms for free filtration [4]. Compromised functions of the GFB and the concomitant increase in albumin filtration are usually masked for prolonged periods of time due to albumin reabsorption by renal proximal tubule cells, mediated by receptor-mediated endocytosis [5]. Once the tubular reabsorptive capacity is saturated, albumin excretion in the urine increases [6]. The commonly used limit for microalbuminuria is uACR > 30 mg/g, but even lower uACR values elevate the risk for cardiovascular diseases and mortality [7, 8]. In addition, limitations of formulas for eGFR might lead to an under-diagnosis of early functional changes in the kidney, thus leading to the delayed initiation of reno-protective therapeutic measures [9]. Therefore, there is apparently the need for new non-invasive diagnostic and prognostic biomarkers for CKD. In this context, extracellular vesicles (EVs) detectable in the urine have gained increasing interest [10].

EVs are defined as particles with a lipid bilayer released from cells and, in the context of the kidney, can be easily recovered from urine samples [11]. Furthermore, EVs carry markers specific for the cells of origin and provide information about the content of the parental cells. Thus, the assessment of changes in EV cargo in combination with the origin of the urinary EVs might generate specific insights into localization, cause and progression of different kidney diseases [12, 13].

According to a previous study in animals [14], changes in the permeability of the GFB lead to the accumulation of serum albumin in the subpodocyte space and the subsequent endocytosis by the podocytes [15]. Part of the endocytosed albumin is degraded in lysosomes, whereas the majority is released into the urinary space via transcytosis as albumin containing EVs [14]. It is assumed that only a small proportion of the vesicles is reabsorbed along the proximal tubule, while the majority is excreted in the urine [15]. Accordingly, albumin and the podocyte-specific protein podocalyxin [10], were detected in EVs isolated from the urine of the animals [14].

In view of these results, we hypothesized that urinary vesicular albumin and podocalyxin can be used as novel diagnostic and prognostic markers for the deterioration of kidney function in humans. So far, there are no measurements of urinary vesicular albumin and podocalyxin and evaluations of their relationship to kidney function markers in a human observational study available. Such markers could be particularly informative in old-aged individuals where the prevalence of albuminuria or low eGFR is higher than in younger individuals. We thus measured urinary vesicular albumin and podocalyxin at baseline and follow-up of a population-based cohort study in the elderly (i.e. age 70-95 year at baseline), characterised these novel kidney biomarkers and evaluated their association with eGFR-based CKD and albuminuria cross-sectionally and longitudinally.

## MATERIALS AND METHODS

### AugUR cohort study description

The German AugUR study (<u>A</u>ltersbezogene <u>U</u>ntersuchungen zur <u>G</u>esundheit der <u>U</u>niversität <u>R</u>egensburg) is a prospective study of the general old-aged population in and around the city of Regensburg, Bavaria. AugUR focuses on chronic diseases and associated risk factors in the population aged 70 to 95 years at baseline. Details on the study were published earlier [16–20]. In brief, 1,133 participants were included in the first AugUR survey between 2013 and 2015. A three year follow-up was conducted between 2016 and 2018 with 733 participants.

The AugUR study was approved by the Ethics Committee of the University of Regensburg, Germany (vote 12-101-0258). The study complies with the 1964 Helsinki declaration and its later amendments. All participants provided informed written consent.

### AugUR study program

General medical examinations at the study centre included blood pressure, height, weight, waist and hip circumference amongst others. Obesity was defined as body mass index (BMI) ≥ 30 kg/m². Systolic and diastolic blood pressures (SBP and DBP) were measured by an automatic device three times after >5 min resting, using the average of the second and third measurements in the analyses. Mean arterial pressure (MAP) was calculated by DBP + ((SBP – DBP)/3).

A questionnaire conducted as in-person interview included information on general chronic diseases, medication intake and lifestyle factors like smoking. Coronary artery disease (CAD) was defined if at least one of the following conditions was reported by the participants: myocardial infarction, percutaneous coronary intervention, or coronary artery bypass surgery. Cardiovascular disease (CVD) was defined as CAD or stroke. Hypertension was defined as blood pressure ≥ 140/90 mmHg or if the individual reported a prior hypertension diagnosis and antihypertensive medication intake [21]. Diabetes was defined as self-reported diagnosis of diabetes or the use of antidiabetic medication [22].

### AugUR biomarker assessment

Non-fasting blood samples were drawn in a sitting position after at least 5 min of resting. Mild venous stasis was applied for a maximum duration of 1 min. Blood was taken using a 21G multifly needle. Midstream urine was sampled.

Biobanked samples (stored at -80°C) were used for laboratory analyses for creatinine, cystatin C, albumin and α1-microglobulin on a Siemens Dimension Vista 1500 (Siemens Healthcare, Erlangen, Germany). Analyses were performed in compliance with the “Guidelines of the German Medical Association for Quality Assurance of Medical Laboratory Tests” (RiLiBäK) at the Central Laboratory of the University Hospital Regensburg, which is accredited in accordance with the standard DIN EN ISO 15189. Serum cystatin C was measured with an immunoassay (assay CYSC, [mg/l]). Creatinine from serum and urine was enzymatically measured (assay ECREA, [mg/dl]). Urine albumin was measured with an immunoassay (assay MALB, [mg/l] with a limit of detection (LoD) of 5 mg/l) and α1-microglobulin (α1M, assay A1MIC, [mg/l], LoD = 7.8 mg/l) with nephelometry. Urinary albumin and α1M were normalized to urinary creatinine, i.e. urinary albumin-to-creatinine-ratio (uACR) and urinary α1M-to-creatinine-ratio (uα1MCR) and expressed in [mg/g].

Chronic Kidney Disease Epidemiology Collaboration (CKD-EPI) 2021 equation [23] was used to derive eGFR [ml/min/1.73m²] from serum creatinine and cystatin C. CKD based on eGFR was defined with values <60 ml/min/1.73m². Incident CKD was defined as eGFR <60 ml/min/1.73m² at follow-up and eGFR >60 ml/min/1.73m² at baseline. Microalbuminuria was defined as uACR 30-300 mg/g and macroalbuminuria as uACR >300 mg/g [1]. Incident albuminuria was defined as no albuminuria at baseline and micro- or macroalbuminuria at follow-up.

### Isolation of EVs from urine

EVs from urine were isolated employing a differential centrifugation protocol. Urine (7 ml), stored at -80°C, was thawed in a water bath at 37°C and samples were homogenized by inverting. The urine was treated with EDTA, 100x protease and phosphatase inhibitor (PI) (Thermo Fisher Scientific, 78446), and PonceauS (PonS) (Sigma Aldrich, P7170), so that EDTA and PI were present in 1x concentration and PonS in a ∼1:115 dilution. Samples were subsequently centrifuged at 3,234 g and 4°C for 20 minutes (Eppendorf 5804R Centrifuge, S-4-72) to remove cells and cell fragments. The resulting pellet was discarded, and 7 ml of supernatant were ultracentrifuged at 329,000 g and 4°C for 1h (Optima L-80 XP ultracentrifuge, Optima LE 80-K ultracentrifuge, Centrikon T-1170 ultracentrifuge, 70.1 Ti rotor, TFT 70.13 rotor). The supernatant was discarded, and the pellet was washed twice with PBS. For this, 7 ml of PBS in combination with 60 µl PonS and/or 1,2 ml of PBS in combination with 10 µl PonS were used. Samples were subsequently centrifuged for 1h at 4°C and 329,000g (Optima L-80 XP ultracentrifuge, Optima LE 80-K ultracentrifuge, Centrikon T-1170 ultracentrifuge, 70.1 Ti rotor, TFT 70.13 rotor) and/or 186,000 g (Optima^TM^ MAX-E ultracentrifuge, TLA-55 rotor). Prior to centrifugation in the Optima^TM^ MAX-E ultracentrifuge pellets were transferred into new cups using 3 x 400 µl PBS. The isolated EV pellet was suspended and transferred into a new tube using 2 x 25 µl of a solution containing PBS and 1x PI. Isolated samples were stored at -80°C until further processing.

### EV sample preparation

For downstream analysis, 15 µl of the resuspended EV volume was preserved. The remaining volume was lysed using 10x RIPA buffer (abcam, ab156034). To support the lysis of EV membranes, samples were vortexed and subsequently shaken at 1400 rpm and 4°C for 20 min.

### Fluorescence microscope

In a pilot study, EVs from one young subject were characterized employing the MemGlow^TM^ 560 probe (Cytoskeleton Inc., MG02-02), a fluorogenic probe that integrates into lipid bilayers [24], in combination with an antibody against the podocyte-specific marker protein podocalyxin. Podocalyxin was detected by abcam Rabbit recombinant monoclonal podocalyxin antibody conjugated to Alexa Fluor® 488 (= PODXL488) (ab208254). 50 µl of vesicle suspension (1:5 diluted) were incubated over night at 4°C and 300 rpm with the primary antibody (diluted 1:50). To wash the vesicles, the volume was brought to 1,2 ml and the suspension was subsequently centrifuged at 186,000 g and 4°C for 1h (Optima^TM^ MAX-E ultracentrifuge, TLA-55 rotor). The resulting pellet was resuspended in 50 µl PBS, incubated with 0.2 µM MemGlow^TM^ at 50 rpm for 30 minutes in the dark. Afterwards the previously described washing step was repeated. The stained EV pellet was resuspended in 10 µl of PBS and 3 µl of solution were applied to a microscope slide. Samples were mounted after drying for a few minutes. To identify the proportion of podocyte-derived vesicles in the total population, the dyed EVs were visualized using a Laser Scanning microscope (LSM710, Zeiss, Jena). Data processing and analysis was performed using the Zeiss Zen lite (ZEN 3.9) software. Vesicles that were stained with MemGlow^TM^, and vesicles stained with both the membrane dye and the antibody, were counted in six equally sized squares, which were randomly positioned in the image. Afterwards the proportion of double-positive (and therefore podocyte-derived) vesicles from the MemGlow^TM^-positive vesicles was calculated (Supplementary Figure 1).

### Quantification of EV-albumin and EV-podocalyxin via ELISA

EV-derived albumin and podocalyxin concentrations were quantified using the commercially available human albumin ELISA kit (abcam, ab227933) and the human podocalyxin ELISA kit (reddot Biotech Inc., RD-PCX-Hu), respectively. Lysed samples were diluted in the buffers included in the kits. Standards were solved and diluted, so that the same concentrations of RIPA, PBS and phosphatase and protease inhibitor and PonS were present in both standards and the samples. Samples were initially diluted 1:20 for the albumin ELISA and 1:10 for the podocalyxin ELISA, samples with concentrations above the standard curve were diluted further. All standards were assayed in duplicate. The protein concentrations of the samples were calculated based on the standard curve. Afterwards, the normalized vesicular albumin and podocalyxin concentrations per mL of urine were calculated (see supplementary information). The 0.5% extreme values at the upper end of the distribution for each EV marker at each point in time were removed.

To determine the intra- and inter-assay coefficient of variability (CV) for both vesicular albumin and podocalyxin, samples were measured in triplicate (n = 26). After the initial determination of vesicular albumin and podocalyxin concentration lysed samples were stored for up to 17 months at -80°C. Subsequently, vesicular albumin and podocalyxin concentrations were determined in triplicate on two different days, one week apart. Here, samples with high, medium and low concentrations of vesicular albumin and podocalyxin were diluted 1:40 and 1:60 for the triplicate measurements, respectively. Samples were also stored at -80°C between triplicate measurements. The intra- and inter-assay coefficient of variability (CV) were calculated based on protein concentration prior to normalization (see supplementary information). Inter-assay CV was calculated between the two triplicate measurements one week apart. Additionally, inter-assay CV was calculated between the two triplicate measurements and initial vesicular albumin and podocalyxin measurement, thus reflecting the stability of vesicular albumin and podocalyxin concentrations over long storage periods.

Since spot urine samples were collected, vesicular albumin and podocalyxin values were normalized to urinary creatinine (termed vACR and vPCR, respectively; see supplementary information). Both, vACR and vPCR were not normally distributed. Therefore, for regression analyses ln-scales values were used and graphic representations were given on log-scale.

### Analysis sample

For the present analysis, we included data from all AugUR participants with available urine biomarkers and eGFR for baseline and three-year follow-up (n=580).

### Statistical analysis

Data management and statistical analyses were performed using SAS 9.4 software (SAS Institute Inc., Cary, NC, USA) and IBM SPSS Statistics for Windows, Version 29.0.2 (IBM Corp., Armonk, NY, USA). Categorical variables were described with absolute numbers and percent. Normally distributed variables were given as mean ± standard deviation (SD). The median and interquartile range were used to describe the distribution of skewed variables. Estimates were given with 95% confidence intervals (CI). Spearman’s correlation coefficient r was reported with 95% CI calculated after r-to-z transformation with estimated standard errors. Linear and logistic regression analyses were performed to obtain estimates and 95% CI for continuous and binary outcomes, respectively. Explained variance in linear regression models was expressed as R². Cox proportional hazard regression analyses were used to obtain hazard ratios (HR) and 95% CI.

## RESULTS

### Study sample characteristics

A total of 580 AugUR participants aged 70 to 95 years at baseline (59% men) were included in the study (Table 1). Median follow-up time was 3.2 years. Kidney function relevant biomarkers were measured at baseline and follow-up.

**Table 1.**
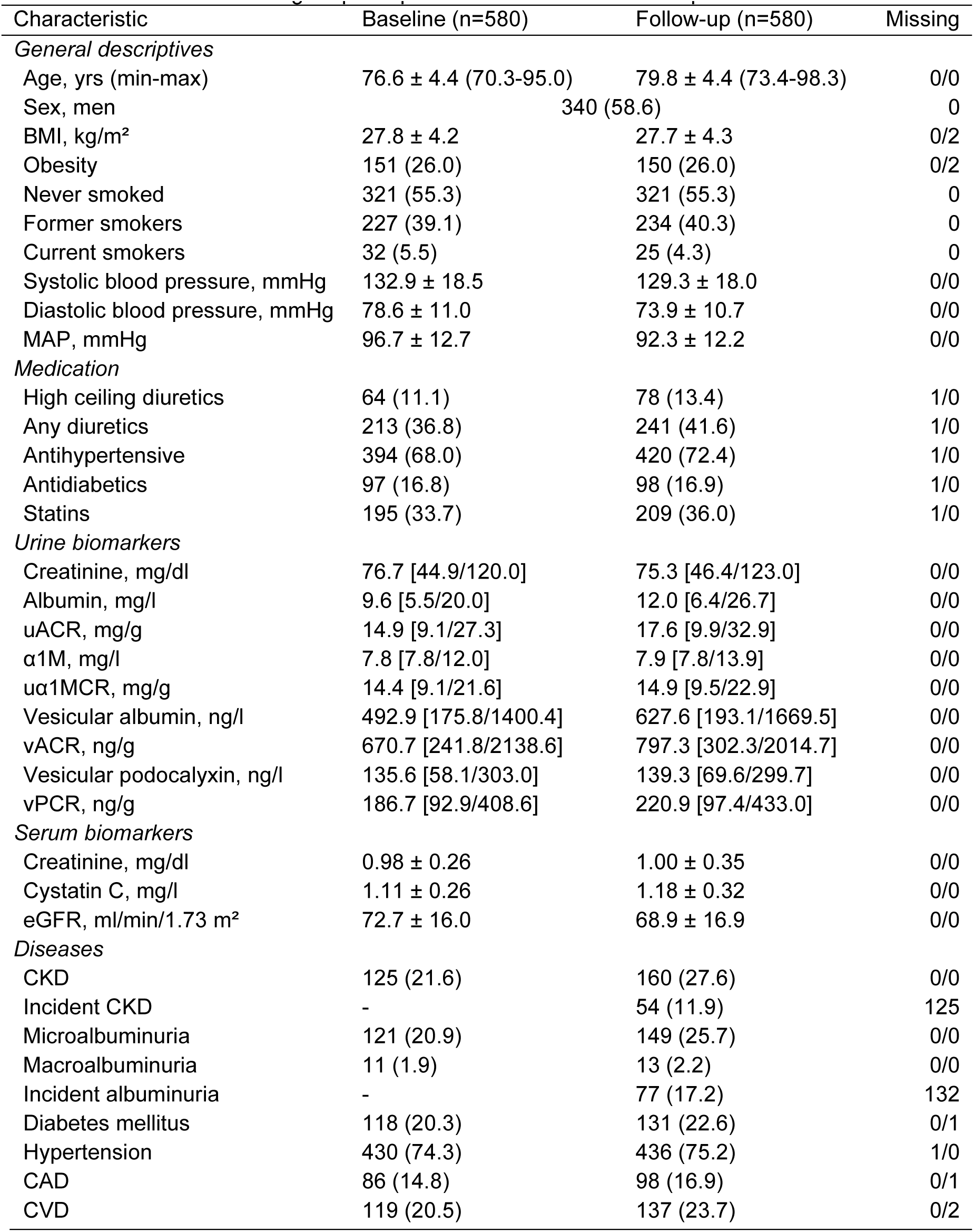
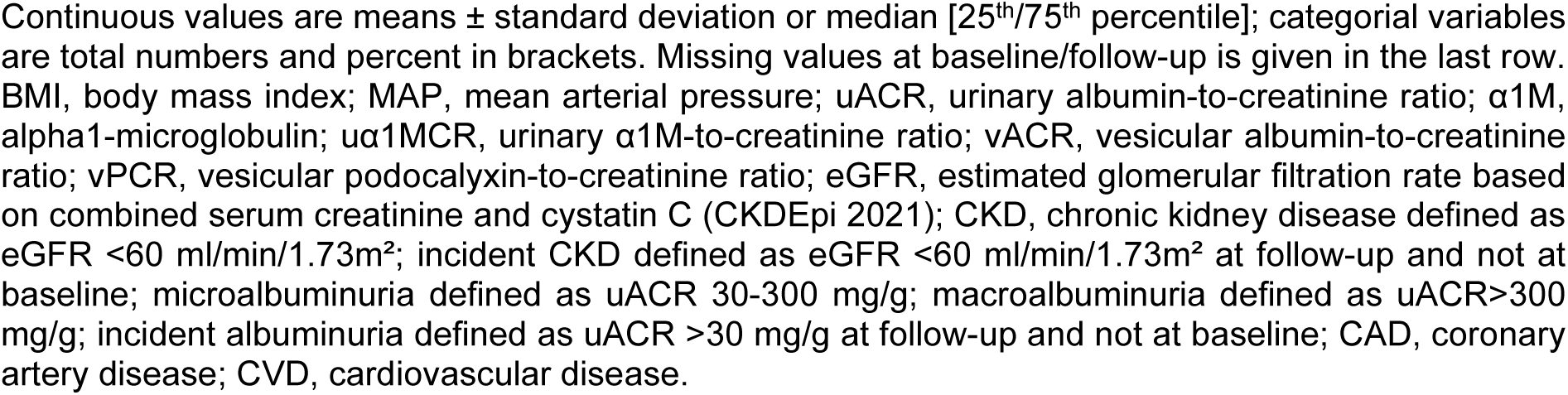
Characteristics of AugUR participants at baseline and follow-up.

| Characteristic | Baseline (n=580) | Follow-up (n=580) | Missing |
| --- | --- | --- | --- |
| *General descriptives* |  |  |  |
| Age, yrs (min-max) | 76.6 ± 4.4 (70.3-95.0) | 79.8 ± 4.4 (73.4-98.3) | 0/0 |
| Sex, men | 340 (58.6) |  | 0 |
| BMI, kg/m<sup>2</sup> | 27.8 ± 4.2 | 27.7 ± 4.3 | 0/2 |
| Obesity | 151 (26.0) | 150 (26.0) | 0/2 |
| Never smoked | 321 (55.3) | 321 (55.3) | 0 |
| Former smokers | 227 (39.1) | 234 (40.3) | 0 |
| Current smokers | 32 (5.5) | 25 (4.3) | 0 |
| Systolic blood pressure, mmHg | 132.9 ± 18.5 | 129.3 ± 18.0 | 0/0 |
| Diastolic blood pressure, mmHg | 78.6 ± 11.0 | 73.9 ± 10.7 | 0/0 |
| MAP, mmHg | 96.7 ± 12.7 | 92.3 ± 12.2 | 0/0 |
| *Medication* |  |  |  |
| High ceiling diuretics | 64 (11.1) | 78 (13.4) | 1/0 |
| Any diuretics | 213 (36.8) | 241 (41.6) | 1/0 |
| Antihypertensive | 394 (68.0) | 420 (72.4) | 1/0 |
| Antidiabetics | 97 (16.8) | 98 (16.9) | 1/0 |
| Statins | 195 (33.7) | 209 (36.0) | 1/0 |
| *Urine biomarkers* |  |  |  |
| Creatinine, mg/dl | 76.7 [44.9/120.0] | 75.3 [46.4/123.0] | 0/0 |
| Albumin, mg/l | 9.6 [5.5/20.0] | 12.0 [6.4/26.7] | 0/0 |
| uACR, mg/g | 14.9 [9.1/27.3] | 17.6 [9.9/32.9] | 0/0 |
| α1M, mg/l | 7.8 [7.8/12.0] | 7.9 [7.8/13.9] | 0/0 |
| uα1MCR, mg/g | 14.4 [9.1/21.6] | 14.9 [9.5/22.9] | 0/0 |
| Vesicular albumin, ng/l | 492.9 [175.8/1400.4] | 627.6 [193.1/1669.5] | 0/0 |
| vACR, ng/g | 670.7 [241.8/2138.6] | 797.3 [302.3/2014.7] | 0/0 |
| Vesicular podocalyxin, ng/l | 135.6 [58.1/303.0] | 139.3 [69.6/299.7] | 0/0 |
| vPCR, ng/g | 186.7 [92.9/408.6] | 220.9 [97.4/433.0] | 0/0 |
| *Serum biomarkers* |  |  |  |
| Creatinine, mg/dl | 0.98 ± 0.26 | 1.00 ± 0.35 | 0/0 |
| Cystatin C, mg/l | 1.11 ± 0.26 | 1.18 ± 0.32 | 0/0 |
| eGFR, ml/min/1.73 m<sup>2</sup> | 72.7 ± 16.0 | 68.9 ± 16.9 | 0/0 |
| *Diseases* |  |  |  |
| CKD | 125 (21.6) | 160 (27.6) | 0/0 |
| Incident CKD | - | 54 (11.9) | 125 |
| Microalbuminuria | 121 (20.9) | 149 (25.7) | 0/0 |
| Macroalbuminuria | 11 (1.9) | 13 (2.2) | 0/0 |
| Incident albuminuria | - | 77 (17.2) | 132 |
| Diabetes mellitus | 118 (20.3) | 131 (22.6) | 0/1 |
| Hypertension | 430 (74.3) | 436 (75.2) | 1/0 |
| CAD | 86 (14.8) | 98 (16.9) | 0/1 |
| CVD | 119 (20.5) | 137 (23.7) | 0/2 |

Continuous values are means ± standard deviation or median [25<sup>th</sup>/75<sup>th</sup> percentile]; categorical variables are total numbers and percent in brackets. Missing values at baseline/follow-up is given in the last row. BMI, body mass index; MAP, mean arterial pressure; uACR, urinary albumin-to-creatinine ratio; α1M, alpha1-microglobulin; uα1MCR, urinary α1M-to-creatinine ratio; vACR, vesicular albumin-to-creatinine ratio; vPCR, vesicular podocalyxin-to-creatinine ratio; eGFR, estimated glomerular filtration rate based on combined serum creatinine and cystatin C (CKDEpi 2021); CKD, chronic kidney disease defined as eGFR <60 ml/min/1.73m<sup>2</sup>; incident CKD defined as eGFR <60 ml/min/1.73m<sup>2</sup> at follow-up and not at baseline; microalbuminuria defined as uACR 30-300 mg/g; macroalbuminuria defined as uACR >300 mg/g; incident albuminuria defined as uACR >30 mg/g at follow-up and not at baseline; CAD, coronary artery disease; CVD, cardiovascular disease.

### Characteristics of vesicular albumin and podocalyxin at baseline

From biobanked urinary samples, extracellular vesicles could be isolated (Supplementary Figure 1). Vesicular albumin and podocalyxin concentrations ranged from 45 to 18,849 ng/l and 11 to 10,499 ng/l, respectively. The stability of the quantitation results for vesicular albumin and podocalyxin in repeat measurements under different storage times was assessed (Supplementary Table 1). The intra-assay coefficient of variability (CV) was below 10% for both markers, whereas the inter-assay CV was higher for vesicular albumin compared to podocalyxin (27% and 13%, respectively).

To correct for urine volume, vesicular albumin and podocalyxin were normalized to urinary creatinine (vACR, vPCR). All further analyses were performed with vACR and vPCR, respectively. At baseline, the range for vACR was 37 ng/g to 42,890 ng/g and 6.4 ng/g to 40,272 ng/g for vPCR. To gain normal distributions for regression analyses, baseline vACR and vPCR were ln-transformed resulting in ranges of 3.6 to 10.7 for vACR and 1.9 to 10.6 for vPCR, respectively.

We analysed the associations of age and sex with vACR and vPCR (Table 2). Whereas linear regression using ln-scaled vACR as outcome revealed no association with age (p=0.671), ln-scaled vPCR was significantly associated with age (b=0.038 per year, 95% CI=0.014 to 0.062, p=0.002, R²=1.6%). We found a strong sex difference for vACR (b= -1.031 on ln-scale for men compared to women, 95% CI= -1.244 to -0.818, p=5.31*10^-20^), explaining 13.5% (R²) of vACR variability at baseline. In contrast, no association between vPCR and sex (p=0.775) was detected. Supplementary Figure 2 shows the baseline distributions of vACR and vPCR between age groups and sex.

**Table 2.**
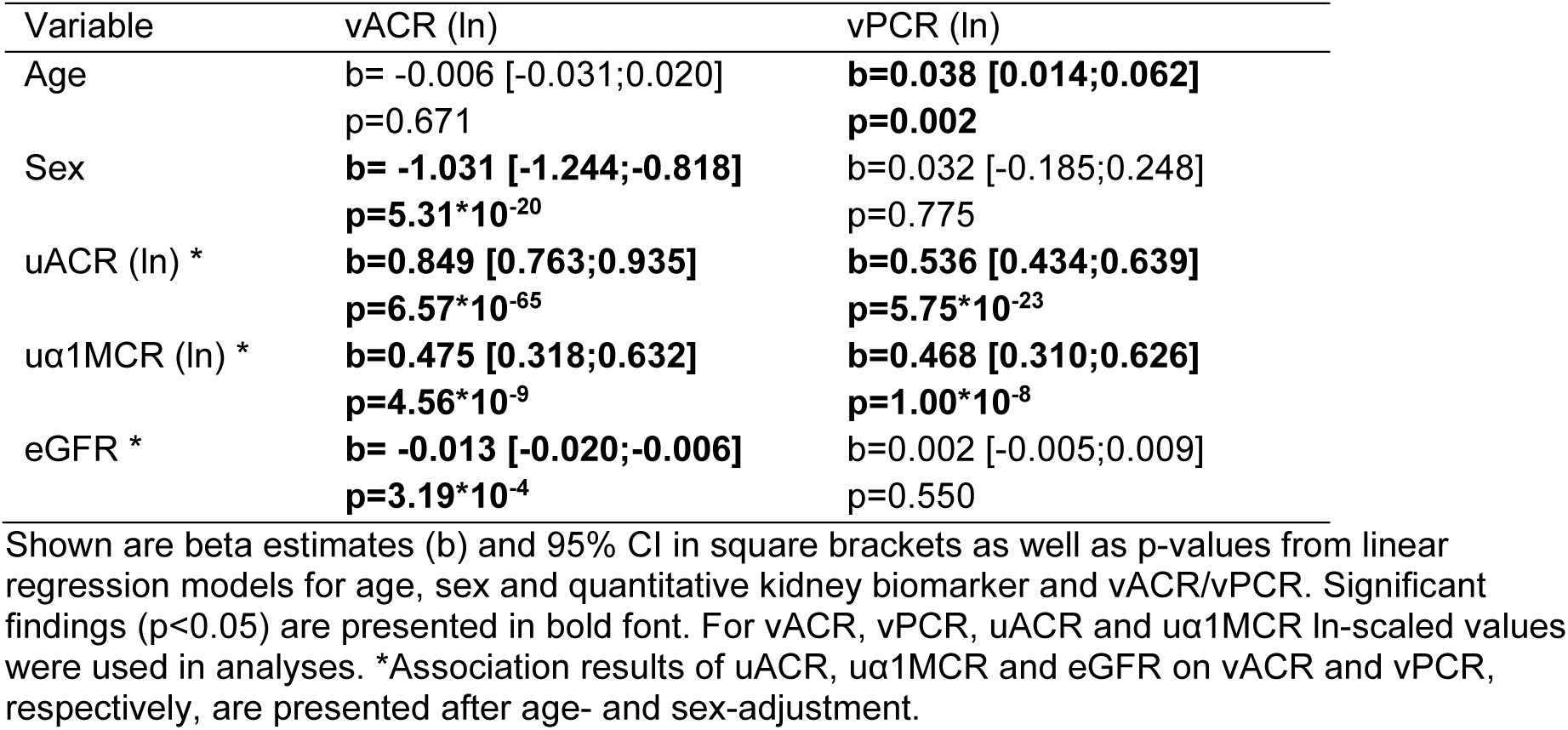
Associations of age, sex and quantitative kidney relevant biomarkers with vACR and vPCR at baseline.

| Variable | vACR (ln) | vPCR (ln) |
| --- | --- | --- |
| Age | b= -0.006 [-0.031;0.020]<br>p=0.671 | **b=0.038 [0.014;0.062]**<br>**p=0.002** |
| Sex | **b= -1.031 [-1.244;-0.818]**<br>**p=5.31\*10<sup>-20</sup>** | b=0.032 [-0.185;0.248]<br>p=0.775 |
| uACR (ln) * | **b=0.849 [0.763;0.935]**<br>**p=6.57\*10<sup>-65</sup>** | **b=0.536 [0.434;0.639]**<br>**p=5.75\*10<sup>-23</sup>** |
| uα1MCR (ln) * | **b=0.475 [0.318;0.632]**<br>**p=4.56\*10<sup>-9</sup>** | **b=0.468 [0.310;0.626]**<br>**p=1.00\*10<sup>-8</sup>** |
| eGFR * | **b= -0.013 [-0.020;-0.006]**<br>**p=3.19\*10<sup>-4</sup>** | b=0.002 [-0.005;0.009]<br>p=0.550 |

Shown are beta estimates (b) and 95% CI in square brackets as well as p-values from linear regression models for age, sex and quantitative kidney biomarker and vACR/vPCR. Significant findings ( $p < 0.05$ ) are presented in bold font. For vACR, vPCR, uACR and uα1MCR ln-scaled values were used in analyses. \*Association results of uACR, uα1MCR and eGFR on vACR and vPCR, respectively, are presented after age- and sex-adjustment.

### Correlations of vesicular albumin and podocalyxin with each other and with kidney function biomarkers at baseline

Vesicular albumin and podocalyxin were moderately correlated (Figure 1), indicating possible independent biological mechanisms of both markers. To evaluate the influence of sex on this correlation, we calculated the explained variance (R²) separately for men and women. R² on ln-scale of vACR and vPCR was 12.7% (7.6% for women and 20.1% for men), possibly resulting from the sex-dependency of vACR.

**Figure 1.**
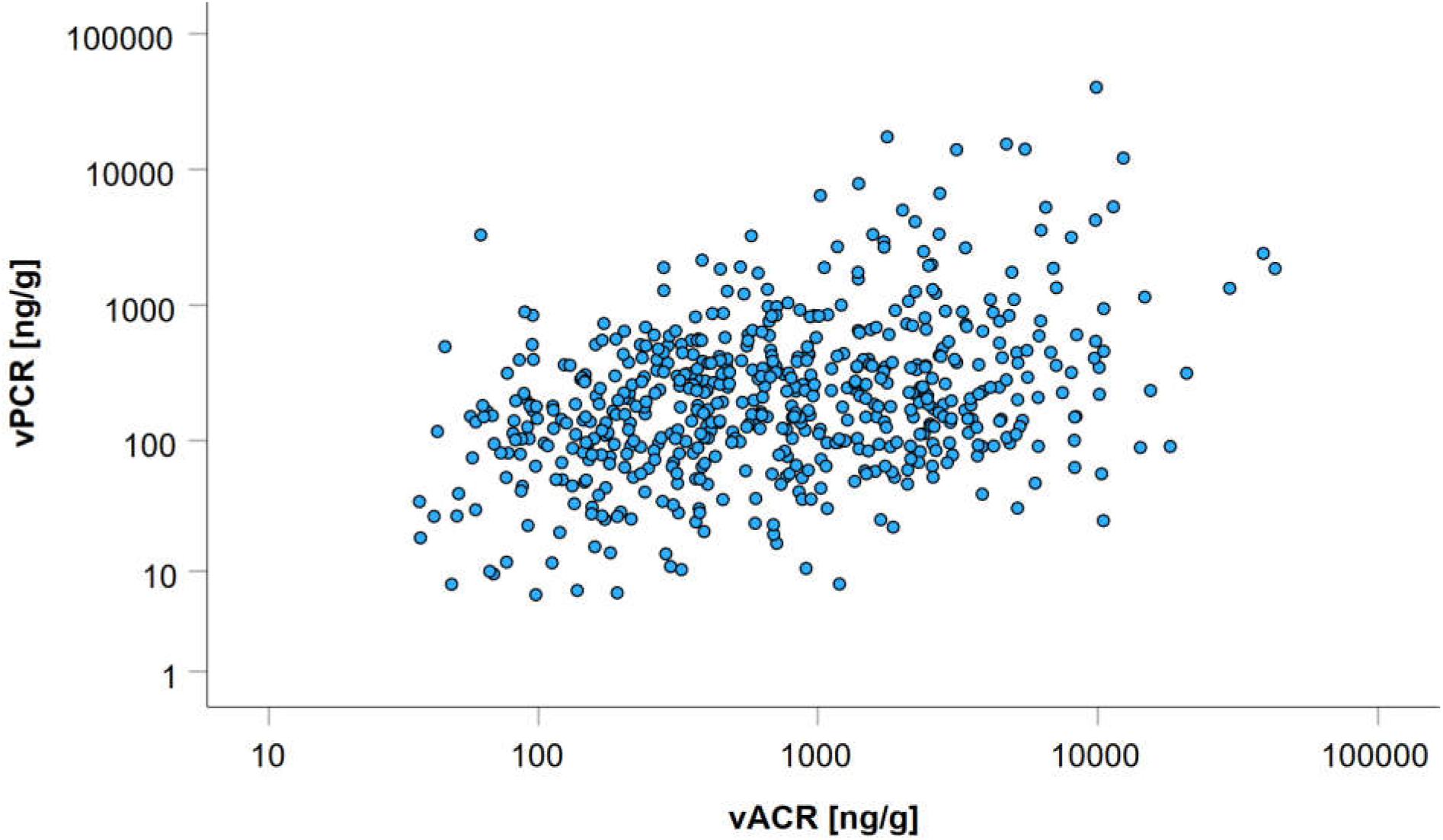
Correlation between vACR and vPCR from 580 urinary samples at baseline. Measurements are given in ng/g and plotted on log scale. Spearman’s correlation between vACR and vPCR r was 0.31 [95% CI=0.23-0.39], p=1.8*10^-14^.

There was no correlation of both vACR and vPCR with eGFR at baseline (Figure 2A, D). Correlations of vACR and vPCR, respectively, with uACR was moderate (Figure 2B, E), and low with uα1MCR (Figure 2C, F).

**Figure 2.**
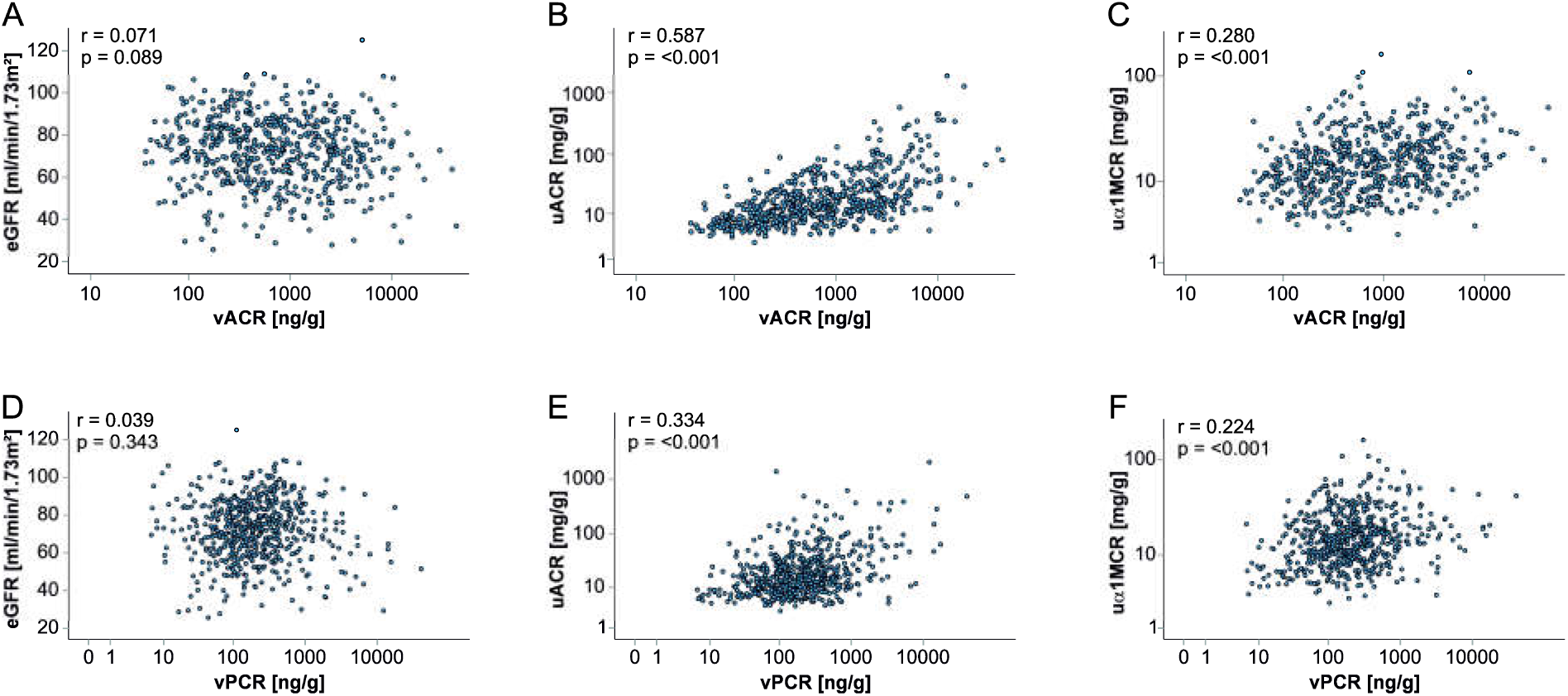
Correlations between vACR, vPCR and kidney markers eGFR, uACR, and uα1MCR at baseline (n=580). On the x-axis vACR (A-C) and vPCR (D-F) are plotted in ng/g on log-scale and on the y-axis (A, D) eGFR in ml/min/1.73 m², (B, E), uACR in mg/g on log-scale, (C, F) uα1MCR in mg/g on log-scale. Spearman’s correlation coefficients r and corresponding p-values are given in the respective plots.

For the correlation of vACR with uACR, Spearman’s correlation coefficient r was 0.59 [95% CI=0.53-0.64] and for the correlation of vPCR with uACR, r was 0.33 [95% CI=0.26-0.41]. R² for vACR and uACR on ln-scale was 22.5% for women and 49.3% for men (36.1 % for both sexes combined). R² for vPCR and uACR on ln-scale was 22.3% for women and 13.8% for men (16.1 % for both sexes combined).

### Association of age, sex and known kidney markers with vesicular albumin and podocalyxin at baseline

Age was associated with vPCR and sex with vACR. To analyse the influence of established kidney biomarkers on vesicular albumin and podocalyxin, we tested the association of uACR, uα1MCR as well as eGFR with vACR and vPCR, respectively, adjusted for age and sex via linear regression at baseline (Table 2). After adjusting for age and sex, uACR was positively associated with vACR as well as vPCR, with stronger effects on vACR. Additionally, uα1MCR, a marker for tubular function, was associated with both vesicular markers in the same direction and with comparable effect sizes. In contrast to vPCR, eGFR was negatively associated with vACR, indicating that higher vACR values were associated with lower eGFR after age- and sex-adjustment. Significant correlation of vACR with eGFR was not observed in unadjusted analyses (Figure 2A). However, the effect of eGFR on vACR was weak (b= -0.013) and seemed to be mainly driven by sex, indicating a sex-eGFR interaction effect on vACR in cross-sectional data.

### Association of baseline vesicular albumin and podocalyxin with incident eGFR-based CKD and incident albuminuria

To assess the potential predictive ability of the two vesicular biomarkers, we restricted to individuals that had (i) no CKD (i.e. eGFR >60 ml/min/1.73 m²) or (ii) no albuminuria (i.e. uACR <30mg/g) at baseline and observed a total of n=54 incident CKD and n=77 incident albuminuria cases. We tested associations of vACR and vPCR at baseline with incident eGFR-based CKD and albuminuria and found significantly elevated risk for eGFR-based CKD with higher vACR levels at baseline in the unadjusted model (HR=1.218, p=0.047) and after adjustment for age and sex (HR=1.268, p=0.021) (Table 3). However, a predictive marker for CKD should be independent of baseline eGFR and uACR. The effect of vACR on incident eGFR-based CKD disappeared when including baseline eGFR and uACR into the model (HR=1.053, p=0.704) (Table 3). In contrast, higher vPCR levels at baseline were associated with lower risk of incident albuminuria after adjustment for eGFR and uACR (HR=0.763, p=0.017). This corresponded with lower vPCR by one unit on ln-scale to confer an HR of 1.3 and thus a 30% increased risk of albuminuria within 3 years (over a range of 8.7 ln-units of vPCR in our data). Additional adjustment for uα1MCR did not change the effect estimate markedly, as well as adding vACR to the model (Table 3). This latter model 6 accounted for a potential confounding effect of vACR on vPCR association with incident albuminuria.

**Table 3.**
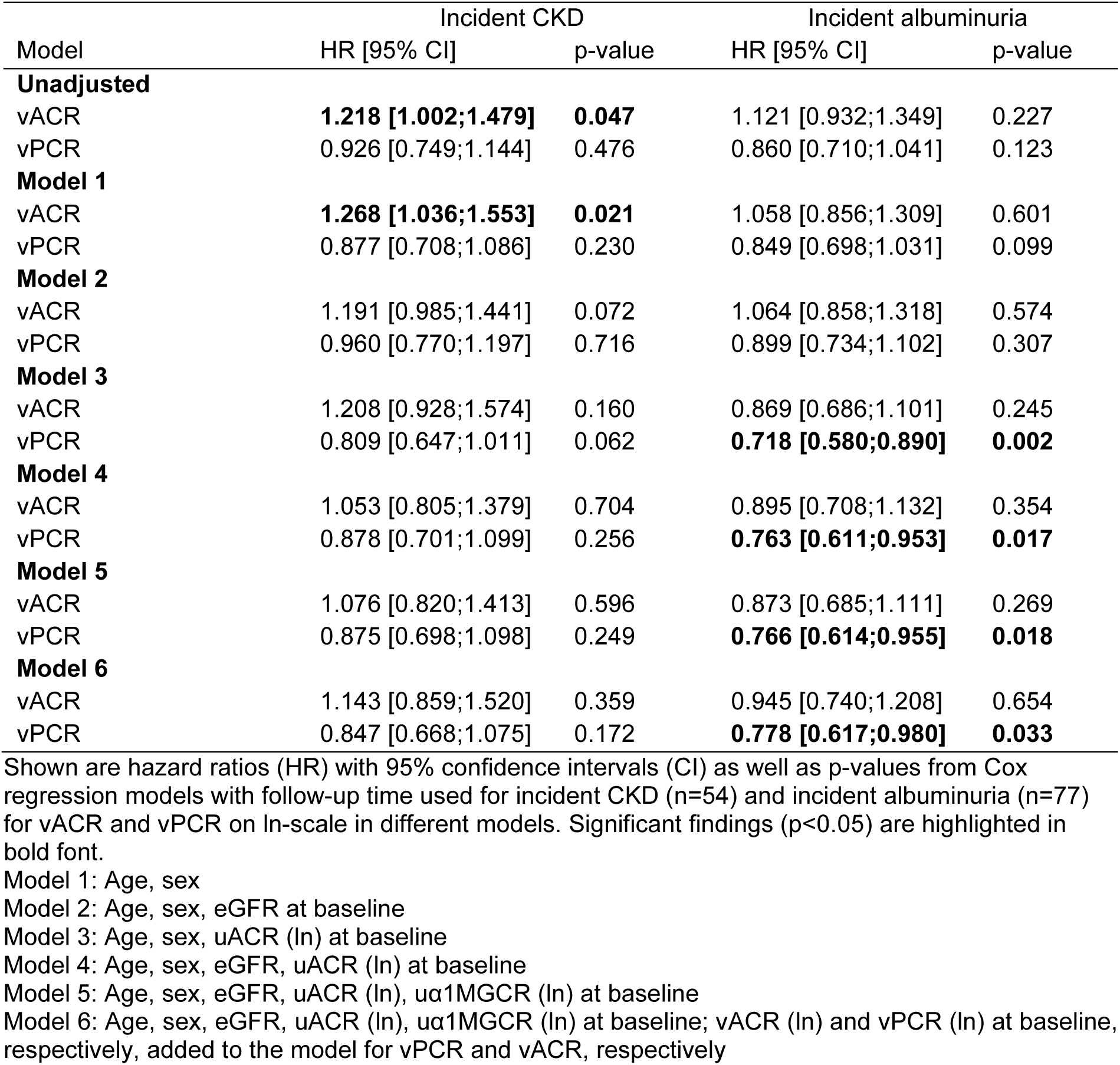
Associations of vACR and vPCR at baseline on ln-scale with incident eGFR-based CKD and incident albuminuria.

In summary, our fully adjusted model indicated a potential predictive power of higher vesicular podocalyxin at baseline for lower incident albuminuria risk.

### Associations of changes in vesicular biomarkers with incident albuminuria and CKD

When analysing vACR and vPCR changes over time, we observed a different pattern regarding incident albuminuria. Across the distribution of vACR at baseline, more participants with higher vACR values at follow-up compared to baseline developed incident albuminuria (n=63) in contrast to those with lower follow-up values (n=14) (Figure 3). Similarly, for vPCR participants with higher values at follow-up compared to baseline developed more often incident albuminuria (n=55) in contrast to those with lower follow-up values (n=22) (Figure 4). An illustration of vACR and vPCR changes between baseline and follow-up over age is given in Supplementary Figure 3.

**Figure 3.**
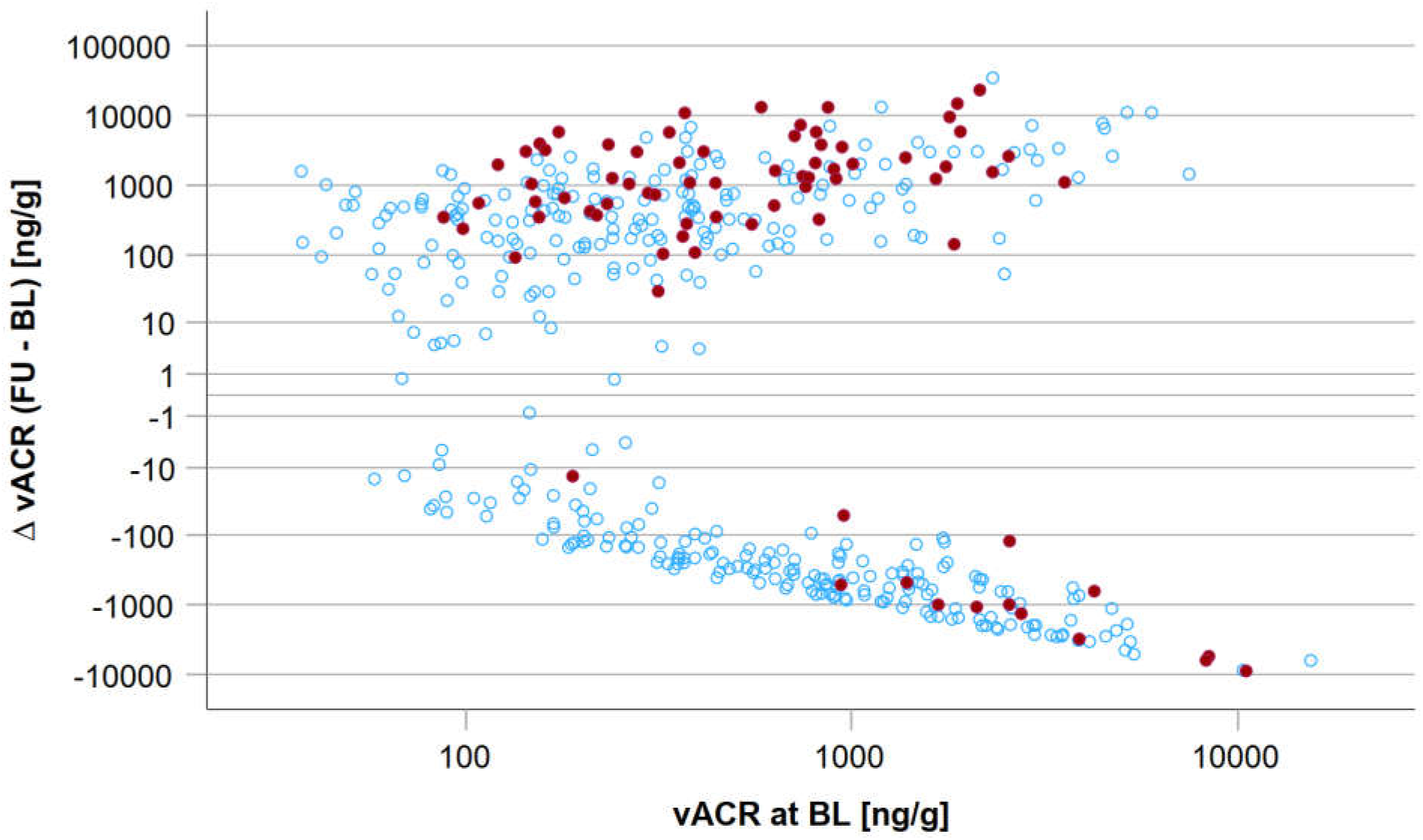
Change in vACR between baseline (BL) and follow-up (FU) on the y-axis (log-scale) over the range of vACR at baseline (x-axis, log-scale). Red dots indicate incident albuminuria cases (n=77) and blue dots participants without incident albuminuria (n=371).

**Figure 4.**
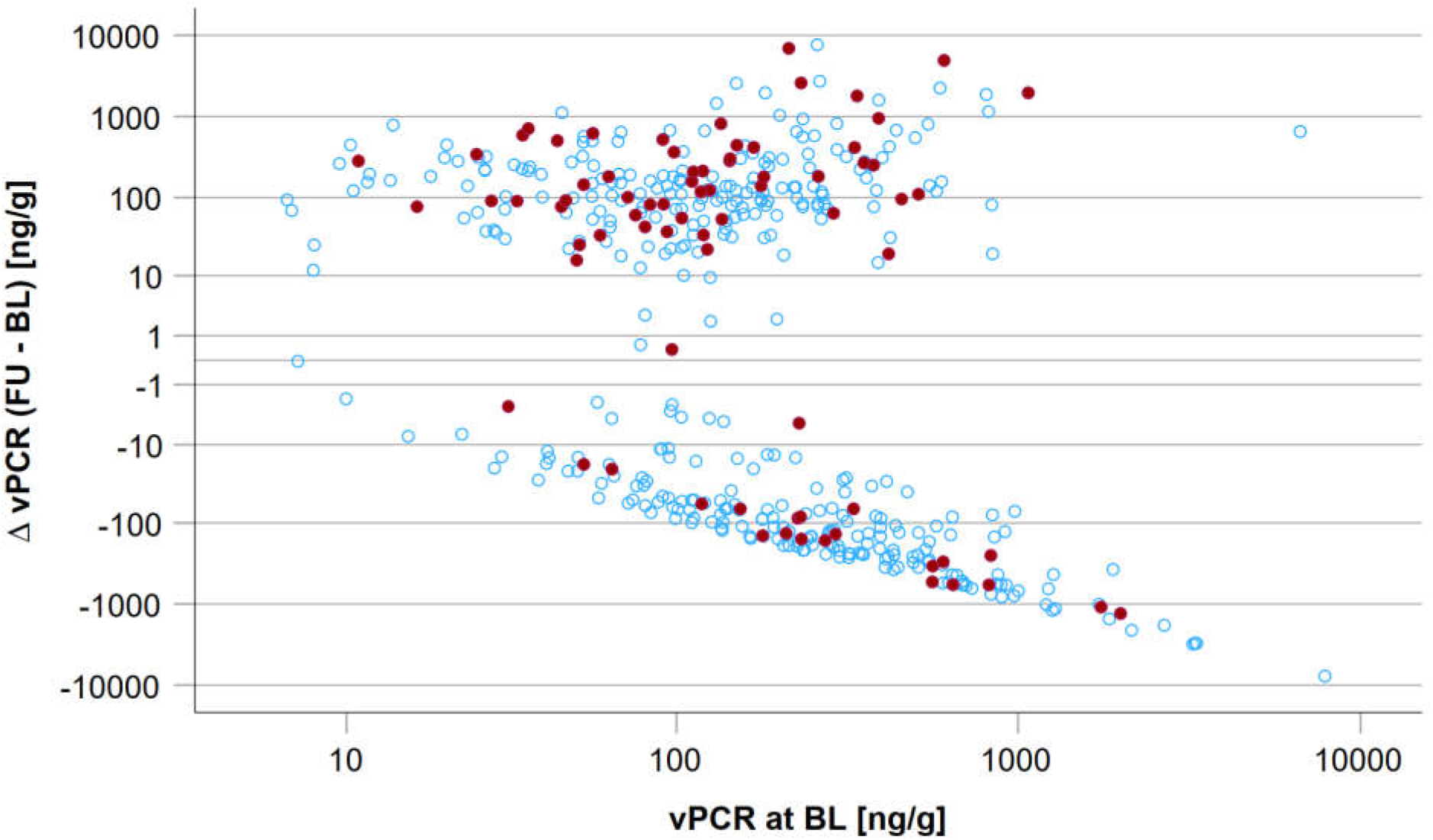
Change in vPCR between baseline (BL) and follow-up (FU) on the y-axis (log-scale) over the range of vPCR at baseline (x-axis, log-scale). Red dots indicate incident albuminuria cases (n=77) and blue dots participants without incident albuminuria (n=371).

For better interpretation, difference of vACR at baseline and follow-up was z-transformed. Difference of vACR at baseline and follow-up was a predictor for incident albuminuria (HR=1.445 [95%CI=1.260; 1.659], p=1.50*10^-7^). After adjustment of age and sex, the association was still significant (HR=1.465 [1.271; 1.688], p=1.29*10^-7^). After additional adjustment for uACR (ln-scale) at baseline, the effect was stronger (HR=1.703 [1.437; 2.019], p=8.09*10^-10^). Additionally adding vACR at baseline (ln-scale) to the model did not alter the effect substantially (HR=1.770 [1.469; 2.133]; p=2.00*10^-9^).

To normalize for baseline vACR values, we further used 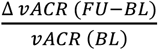. We defined vACR change as ‘stable’ in the range of -1 to 1 after normalization on vACR at baseline (n=393) and ‘increase’ with values >1 (n=187). Figure 5 shows incident albuminuria cases at follow-up over the distribution of 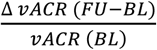

**Figure 5.**
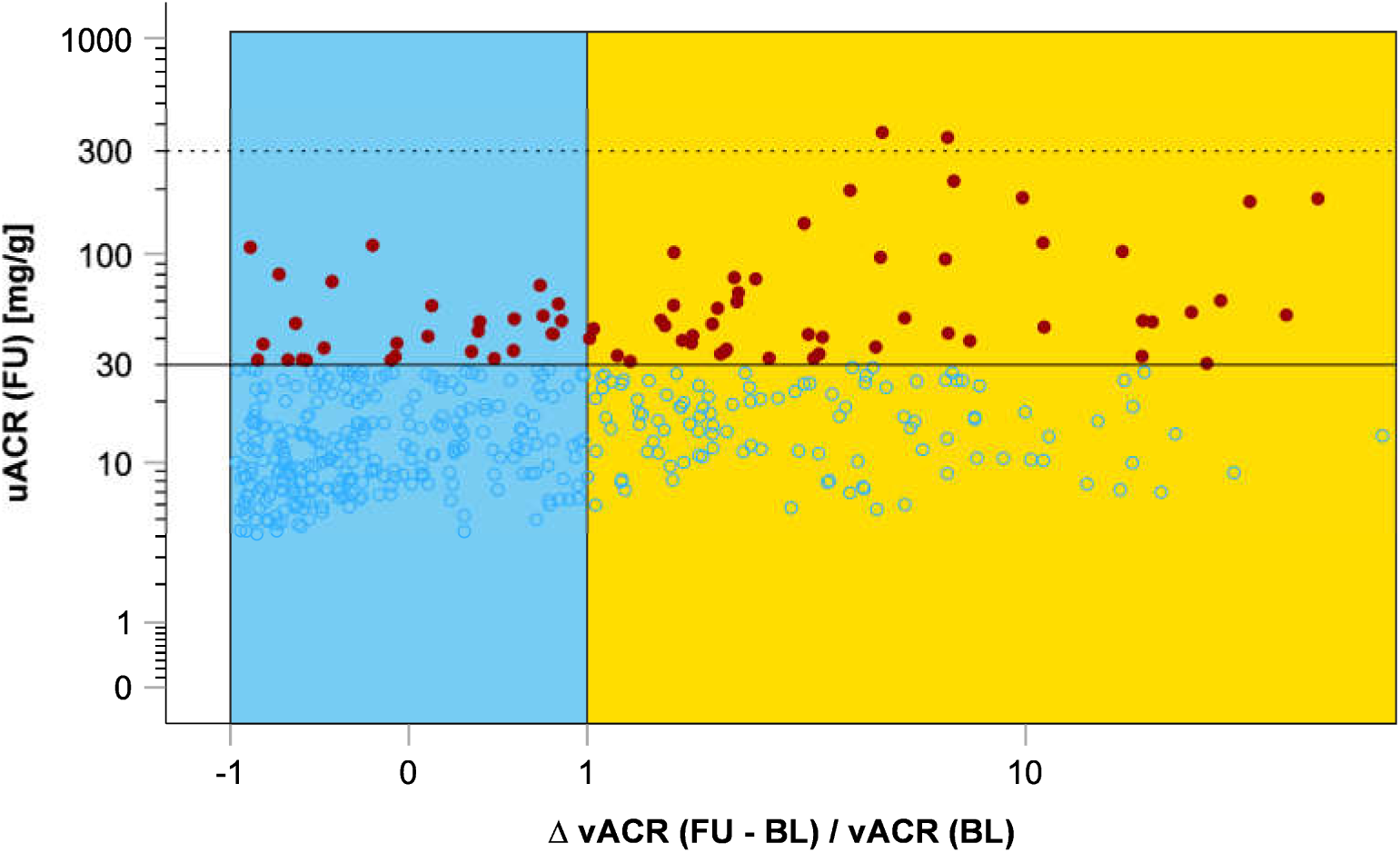
Change of vACR between baseline (BL) and follow-up (FU) after normalization for baseline vACR and occurrence of incident albuminuria. On the y-axis follow-up uACR [mg/g] levels are plotted on log scale with solid line indicating 30 mg/g (microalbuminuria) and dashed line for 300 mg/g (macroalbuminuria). Red dots mark incident albuminuria events (n=77; n=75 incident microalbuminuria, n=2 incident macroalbuminuria) in participants without albuminuria at baseline (n=371). On the x-axis the quotient of the vACR follow-up to baseline difference and baseline vACR is plotted on log scale. Stable quotient was defined as values between -1 and +1 (blue, n=291), whereas increase was defined as quotient > 1 (orange, n=157). In the vACR stable region, 9.6% incident albuminuria cases (n=28) occurred, whereas 31.2% incident albuminuria cases (n=49) were observed in the vACR increase region.

Z-transformation was also computed for vPCR. Difference of vPCR at baseline and follow-up was not a predictor for incident albuminuria (HR=1.141 [95%CI=0.833; 1.562], p=0.411). After adjustment for age and sex, the association was still not significant (HR=1.127 [0.809; 1.570], p=0.481). Additional adjustment for uACR (ln-scale) at baseline, did not alter the effect (HR=1.115 [0.792; 1.572], p=0.532). Taking additionally vPCR at baseline (ln-scale) into the model did not alter the effect substantially (HR=1.083 [0.736; 1.592]; p=0.686).

As for vACR, to normalize for baseline vPCR, we further used 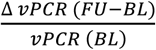. We defined vPCR change as ‘stable’ in the range of -1 to 1 after normalization on vPCR at baseline (n=426) and ‘increase’ with values >1 (n=154). Figure 6 shows incident albuminuria cases at follow-up over the distribution of 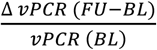

**Figure 6.**
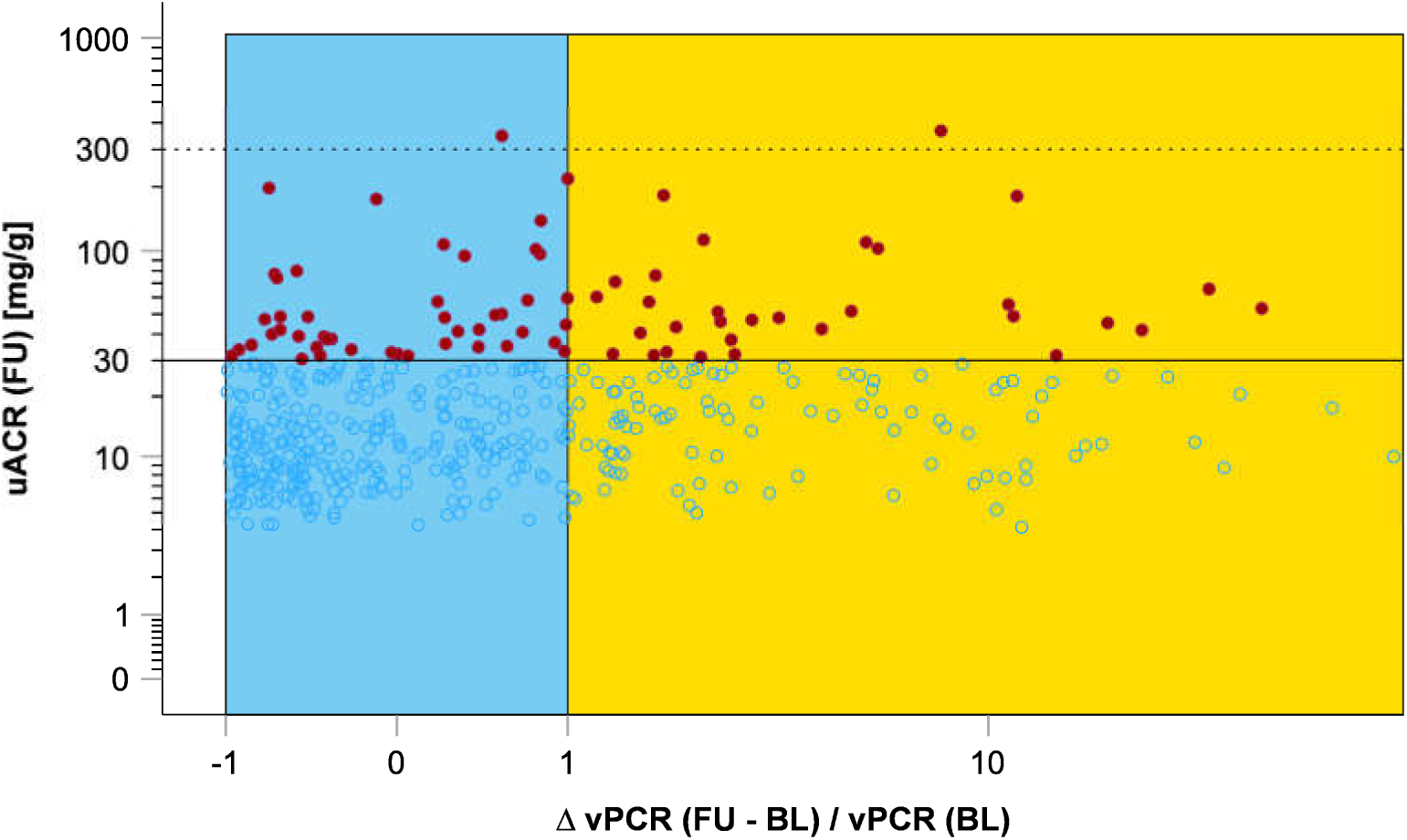
Change of vPCR between baseline (BL) and follow-up (FU) after normalization for baseline vPCR and occurrence of incident albuminuria. On the y-axis follow-up uACR [mg/g] levels are plotted on log scale with solid line indicating 30 mg/g (microalbuminuria) and dashed line for 300 mg/g (macroalbuminuria). Red dots mark incident albuminuria events (n=77; n=75 incident microalbuminuria, n=2 incident macroalbuminuria) in participants without albuminuria at baseline (n=371). On the x-axis the quotient of the vPCR follow-up to baseline difference and baseline vPCR is plotted on log scale. Stable quotient was defined as values between -1 and +1 (blue, n=317), whereas increase was defined as quotient > 1 (orange, n=131). In the vPCR stable region, 14.5% incident albuminuria cases (n=46) occurred, and 23.7% incident albuminuria cases (n=31) were observed in the vPCR increase region.

Change in vPCR shows less pronounced pattern of incident albuminuria distribution over the distribution of 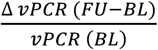 (Figure 6).

The dichotomized variables (stable between follow-up and baseline versus higher values in follow-up) of 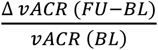 and 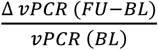 were tested for association with incident albuminuria and CKD (Table 4). Both, vACR- and vPCR-based analyses showed associations with incident albuminuria, but not with CKD. Participants with increasing vACR and vPCR levels over time showed an increased risk for incident albuminuria.

**Table 4.** Associations of dichotomized 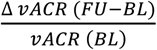 and 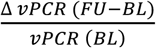 with incident eGFR-based CKD and incident albuminuria.

|  | Incident CKD |  | Incident albuminuria |  |
| --- | --- | --- | --- | --- |
| Model | OR [95% CI] | p-value | OR [95% CI] | p-value |
| **Unadjusted** |  |  |  |  |
| vACR change | 1.003 [0.544; 1.849] | 0.993 | **4.262 [2.545; 7.136]** | **3.56\*10<sup>-8</sup>** |
| vPCR change | 0.621 [0.309; 1.246] | 0.180 | **1.826 [1.097; 3.041]** | **0.021** |
| **Model 1** |  |  |  |  |
| vACR change | 1.028 [0.553; 1.913] | 0.929 | **4.417 [2.618; 7.453]** | **2.60\*10<sup>-8</sup>** |
| vPCR change | 0.667 [0.329; 1.351] | 0.261 | **1.932 [1.152; 3.240]** | **0.013** |
| **Model 2** |  |  |  |  |
| vACR change | 0.976 [0.500; 1.905] | 0.976 | **4.315 [2.550; 7.304]** | **5.16\*10<sup>-8</sup>** |
| vPCR change | 0.578 [0.273; 1.226] | 0.153 | **1.969 [1.169; 3.319]** | **0.011** |
| **Model 3** |  |  |  |  |
| vACR change | 1.149 [0.610; 2.162] | 0.668 | **5.585 [3.155; 9.886]** | **3.55\*10<sup>-9</sup>** |
| vPCR change | 0.699 [0.343; 1.424] | 0.324 | **2.004 [1.163; 3.454]** | **0.012** |
| **Model 4** |  |  |  |  |
| vACR change | 1.160 [0.583; 2.309] | 0.627 | **5.453 [3.064; 9.704]** | **8.11\*10<sup>-9</sup>** |
| vPCR change | 0.606 [0.283; 1.295] | 0.196 | **2.076 [1.195; 3.607]** | **0.009** |
| **Model 5** |  |  |  |  |
| vACR change | 1.158 [0.581; 2.307] | 0.677 | **5.464 [3.070; 9.723]** | **7.75\*10<sup>-9</sup>** |
| vPCR change | 0.620 [0.288; 1.333] | 0.221 | **2.064 [1.185; 3.595]** | **0.010** |
| **Model 6** |  |  |  |  |
| vACR change | 1.161 [0.583; 2.313] | 0.671 | **5.108 [2.850; 9.156]** | **4.31\*10<sup>-8</sup>** |
| vPCR change | 0.656 [0.301; 1.429] | 0.288 | **1.824 [1.027; 3.238]** | **0.040** |

Shown are the results from logistic regression models for the association of dichotomized $\frac{\Delta vACR (FU-BL)}{vACR (BL)}$
(vACR change) and $\frac{\Delta vPCR (FU-BL)}{vPCR (BL)}$ (vPCR change) with incident CKD (n=54) and incident albuminuria (n=77), respectively, in different models (odds ratio (OR) with 95% confidence intervals (CI) for stable versus increased). Significant findings (p<0.05) are highlighted in bold font.
FU, follow-up; BL, baseline.
Model 1: Age, sex
Model 2: Age, sex, eGFR at baseline
Model 3: Age, sex, uACR (ln) at baseline
Model 4: Age, sex, eGFR, uACR (ln) at baseline
Model 5: Age, sex, eGFR, uACR (ln), uα1MGCR (ln) at baseline
Model 6: Age, sex, eGFR, uACR (ln), uα1MGCR (ln) at baseline; simple vACR and vPCR change (follow-up minus baseline), respectively, added to the model for vPCR and vACR change, respectively

In summary, baseline-normalized changes of both vesicular markers, albumin and podocalyxin, between baseline and follow-up were associated with incident albuminuria but not with incident CKD. After dichotomization, increased versus stable levels of vesicular albumin and podocalyxin were associated with incident albuminuria independently of established kidney function biomarkers (Table 4). Effect of vesicular albumin on incident albuminuria was higher (OR=5.5) compared to podocalyxin (OR=2.1) in the fully adjusted model 5. Model 6 with accounting for the respective other vesicular marker did not markedly alter the associations of both vACR and vPCR change with incident albuminuria.

## DISCUSSION

CKD is defined by the progressive loss of kidney function, as determined by a decline in GFR below and an increase in urinary albumin excretion over a threshold for at least three months [1, 25]. As a drawback, the sensitivity of both parameters is limited, and changes in kidney function remain frequently undetected in the early phase of the course of the disease [26]. Consequently, patients would benefit from novel diagnostic and prognostic biomarkers of kidney function to overcome the limitations of the established markers, such as eGFR and albuminuria. Early biomarkers of kidney disease may also facilitate the initiation of reno-protective measures in a timely manner.

The potential of the RNA and protein content, as well as changed concentration levels of podocyte-derived vesicles as biomarkers of podocyte injury, glomerulonephritis and other kidney diseases has been suggested in several studies [27]. Constituents of podocyte-derived vesicles may serve as biomarkers for kidney disease of various aetiology [28, 29], childhood nephrotic syndrome [30], diabetic nephropathy [31], disease activity in systemic lupus erythematosus [32], and renovascular hypertension [33]. Podocyte-derived vesicles were shown to contain e.g. *CD2AP* and *WT1* mRNA as well as WT1 protein [30, 31, 34]. Levels of urinary exosomal *WT1* mRNA have been found to predict eGFR decline in patients with diabetic nephropathy [34]. Furthermore, the content of podocyte-derived vesicles facilitated differentiating between focal segmental glomerulosclerosis and steroid-sensitive nephrotic syndrome [35]. In addition to the diagnosis of kidney disease, the prognostic potential of podocalyxin-positive vesicles has been addressed before. Miller et al. observed that a model including podocalyxin-positive exosomes was able to predict the development of acute kidney injury after cardiac surgery [36]. Furthermore, urinary podocalyxin concentration was elevated in diabetic patients, and patients with various glomerular diseases in comparison to normal controls, thus indicating its potential as a biomarker for diabetic nephropathy and other glomerular diseases [37].

In this study, we assessed the potential of two vesicular proteins as prognostic markers of kidney function. Vesicular podocalyxin and vesicular albumin are known to be derived from podocytes [10, 14], which functionally and structurally contribute to the integrity of the glomerular filtration barrier of the kidney [15]. We found that about 19% of urinary vesicles were podocyte-derived (Supplementary Figure 1). Additionally, the endocytotic activity of podocytes is assumed to play a role in the clearing of the glomerular filtration barrier from accumulating proteins. Thus, an impaired or saturated clearance mechanism might increase the probability for glomerular injuries [38, 39].

Our analyses in an old-aged cohort showed an association between changes in vACR and vPCR with newly occurring albuminuria, suggesting that increasing levels of vesicular albumin and podocalyxin may be indicative of declined kidney function based on the increasing urinary albumin excretion (uACR). Conversely, no such association was found for changes in eGFR, suggesting that the changes in vACR and vPCR over time are more sensitive to changes in uACR than in eGFR. In conclusion, the presence of albumin-containing podocyte-derived vesicles is an early marker of podocyte injury, whereas eGFR rather reflects renal function decline in the advanced stages of kidney disease [40], thus no association between the vesicular markers and eGFR could be observed within this time frame.

Interestingly, we found an association between higher levels of vPCR at baseline and a reduced risk for incident albuminuria in the models adjusting for baseline uACR, eGFR and uα1MGCR suggesting that the association may be podocyte-specific. The association between higher levels of vPCR and a reduced risk for incident albuminuria may also be related to a higher number of viable and endocytic-active podocytes. Consequently, the clearance of the filtration barrier from accumulating proteins, in particular in the subpodocyte space, may be protective for the function of the GFB [38].

In contrast, higher vACR and also vPCR levels in the follow-up compared to baseline were associated with an increased risk for incident albuminuria. The formation of albumin-containing podocyte-derived vesicles may indicate an increasing number of accumulating proteins in the subpodocyte space, resulting in saturation of the clearance mechanism. This may lead to the deterioration of the integrity of the glomerular filter, thus explaining the increased risk of albuminuria. Furthermore, albumin exposure at high concentrations has been shown to elicit apoptosis and injury in podocytes [41–45]. In addition albumin overload induced morphological changes in podocytes, e.g. a disrupted cytoskeleton [42, 44, 45], which plays a crucial role in normal podocyte structure and function. Disruption of the cytoskeleton might lead to podocyte foot process effacement and subsequently to albuminuria/proteinuria [45, 46].

The present study showed a marked sex difference in vACR, being significantly lower in men compared to women, whereas vPCR did not differ between the sexes. The difference in the sex-dependency of vACR and vPCR suggest that both markers are influenced by independent biological mechanisms. Podocalyxin is a highly specific marker of podocytes [10]. For vesicular albumin, the situation is more complex. Vesicular albumin is generated via cellular uptake and transcytosis by podocytes, as shown in animal studies [14]. Furthermore, vesicular albumin may also be part of the vesicular protein corona [47–51], which has been demonstrated to form spontaneously around the surface of vesicles in biological fluids [52]. The latter would occur independently of podocytes and is presumably dependent on the competition between albumin and other proteins for vesicular binding site. Consequently, differences in the global protein status between men and women may be causal for the observed sex differences in vesicular albumin.

In our longitudinal analyses, we have identified subjects with increased vACR and vPCR levels in the follow-up compared to baseline, but also a high proportion of participants with stable or even declining levels. For both markers, in the group with increased levels, the chance for albuminuria was significantly higher compared to subjects with stable parameters. Therefore, the temporal trajectories in vesicular biomarkers indicate (patho-)physiological changes, as described in more detail above, that increase the risk of albuminuria. We also found that considering baseline levels of the vesicular biomarkers and of uACR, improved the association with incident albuminuria. Therefore, the state of the kidney, especially the glomerulus, at baseline might be a modifying factor of the observed associations.

### Strengths and limitations of the study

We acknowledge some limitations of the current study. First, the AugUR population does not represent the general population aged 70+, since based on the requirement for visits at the study centre, there presumably is some selection for mobile and healthy elderly persons. Therefore, the data may not represent the entire aged population, and samples from subjects with a higher degree of comorbidities and age-related degenerative processes are likely underrepresented.

Second, we observed that applying our EV isolation protocol, 19% of urinary EVs were podocalyxin-positive, and, consequently, of podocyte origin. Thus, as expected, a heterogenous population of various vesicle subtypes, originating from different cell types of the urogenital tract, was isolated employing an ultracentrifugation protocol without additional affinity-based purification [53, 54]. The percentage of podocyte-derived EVs was in a similar range as reported previously, e.g. 11.4 ± 6.4% for patients suffering from renovascular hypertension and 6.8 ± 3,4 % for healthy subjects [33]. Similarly, 23.3% of all EVs isolated by the use of a sucrose gradient were of glomerular origin [55]. Nevertheless, as vesicular albumin and podocalyxin, which are highly podocyte-specific, were determined as potential markers of kidney function, the background caused by non-glomerular EVs likely is of limited relevance. The strength of our study is the analysis of a large number of samples of an old-aged population, with an increased incidence of apparently and inapparently compromised kidney function and an increased risk of the development of kidney disease compared to the normal population. Consequently, we analysed samples from a population that is particularly suited for the assessment of diagnostic and prognostic biomarkers of changes in kidney function.

In summary, we identified higher level of baseline vesicular podocalyxin as a predictor for reduced risk of incident albuminuria. In contrast, we found strong association between increase in vesicular albumin and podocalyxin over time with higher risk for albuminuria. Increasing vACR and vPCR levels were associated with 5.5- and 2-fold higher risk for newly occurring albuminuria compared to stable levels of vACR and vPCR, respectively. Based on our results, baseline uACR levels even below 30 mg/g should be considered for risk prediction models, since they enhance the effect of vACR and vPCR changes on albuminuria. No association between changes in vACR and vPCR levels and newly occurring eGFR-based CKD was detected, suggesting that eGFR is not predominantly determined by podocyte-specific effects, whereas uACR is dependent on the integrity of podocytes.

Further studies with longer follow-ups are needed to elucidate the effect of change in vACR and vPCR with incident albuminuria. The aim of those studies should be to investigate the predictive power of differences of vACR and vPCR between two time points for future development of albuminuria.

Furthermore, the present study was focused on vesicular albumin and podocalyxin as novel diagnostic and prognostic biomarkers of kidney function. The content of podocyte-derived vesicles, however, contains an array of further podocyte-specific markers, some of which may be indicative of changes in podocyte function, and, more general, may provide information about the integrity of the glomerular filtration barrier of the kidney. These parameters should be addressed in future studies.

## STATEMENTS AND DECLARATIONS

### Consent to publish

Not applicable.

### Ethics approval

This study was performed in line with the principles of the Declaration of Helsinki. Approval was granted by the Institutional Review Board of the University of Regensburg (IRB number: 12-101-0258).

### Consent statement

Written informed consent was obtained from all individual participants included in the study.

### Availability of data and materials

The individual data generated and analysed during the current study are not publicly available due to data privacy of study participants. Summary statistics are available from the corresponding author on request.

## Funding

The AugUR study was supported by grants from the German Federal Ministry of Education and Research (BMBF 01ER1206 and BMBF 01ER1507) to I.M.H., by the German Research Foundation (DFG HE 3690/7-1 and BR 6028/2-1) to I.M.H. and C.B. and by institutional budget (University of Regensburg). The project was funded by grants from the German Research Foundation (TRR 374 project-ID 50914993) to I.H.M (TRR 374 projects B2, C6, and INF) and to H.C. (TRR 374 project B2).

## Conflict of interest

Author I.M.H. has received support from Roche Diagnostics for a biomarker project in the AugUR study, but unrelated to the work presented here.

## Authorś Contributions

All authors have contributed to interpreting results and manuscript writing. All authors have read and approved the manuscript. Further contributions are:

L.S.: laboratory measurements, data analysis, statistical analysis, manuscript writing

H.C.dH.: data analysis, statistical analysis, manuscript writing L.M.: laboratory measurements, manuscript writing

R.F.: laboratory measurements, manuscript writing

M.E.Z.: data management, data analysis, manuscript writing

C.B.: study physician, overall medical program study, manuscript writing

I.M.H.: project PI, study PI, project supervision, manuscript design, manuscript writing H.C.: project initiation, project PI, project supervision, manuscript writing

K.J.S.: study coordination, project supervision, data management, statistical analysis, manuscript design, manuscript writing

## Acknowledgements

The authors greatly appreciate the outstanding and committed study assistance of Lydia Mayerhofer, Magdalena Scharl, Sabine Schelter and Josef Simon. We would like to express our special thanks to the study participants for contributing to the AugUR study. The authors also thank Bernhard Gess and Katharina Fremter for their technical support in the lab.

## Supplementary information

*Formula 1a. Normalization of vesicular albumin to suspension volume and volume of urine*

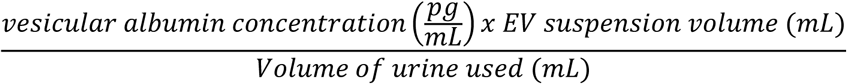

*Formula 1b. Normalization of vesicular podocalyxin to suspension volume and volume of urine*

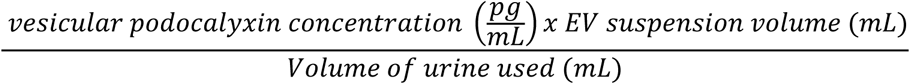

*Formula 2a. Normalization of vesicular albumin concentration on urinary creatinine concentration*

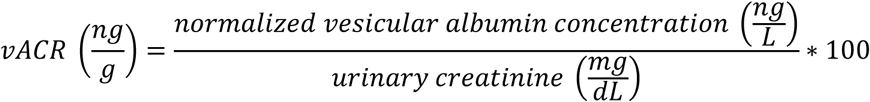

*Formula 2b. Normalization of vesicular podocalyxin concentration on urinary creatinine concentration*

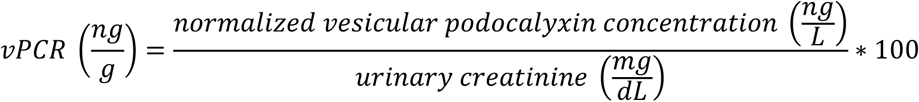

*Formula 3a. Calculation of intra-assay coefficient of variation (CV)*

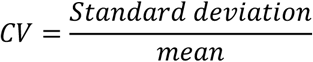

*Formula 3b. Calculation of inter-assay coefficient of variation (CV)*

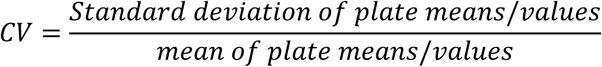

Vesicular albumin and podocalyxin concentrations were determined on two different days a week apart in triplicate (n = 26). Intra- and inter-assay CVs were calculated and the mean, as well as the minimum and maximum of the individual CVs is reported in Supplementary Table 1. After the initial single determination of vesicular albumin and podocalyxin concentrations lysed samples were stored for up to 17 months at -80°C before concentrations of the two markers were determined again as part of the triplicate measurements. Inter-assay CV was also calculated between the two triplicate measurements and the initial vesicular albumin and podocalyxin measurement.

### Isolation of urinary vesicles and quantification of vesicular albumin and podocalyxin

In the pilot study from one sample, 1403 vesicles were counted from which 269 were podocalyxin-positive. Based on this first estimation 19.1% of urinary vesicles seem to be derived from podocytes. Furthermore, these results lead to the assumption that podocalyxin is part of the vesicle membrane, as the antibody binds without prior permeabilization of the vesicle membrane (Supplementary Figure 1).

---

**Supplementary Figure 1.**
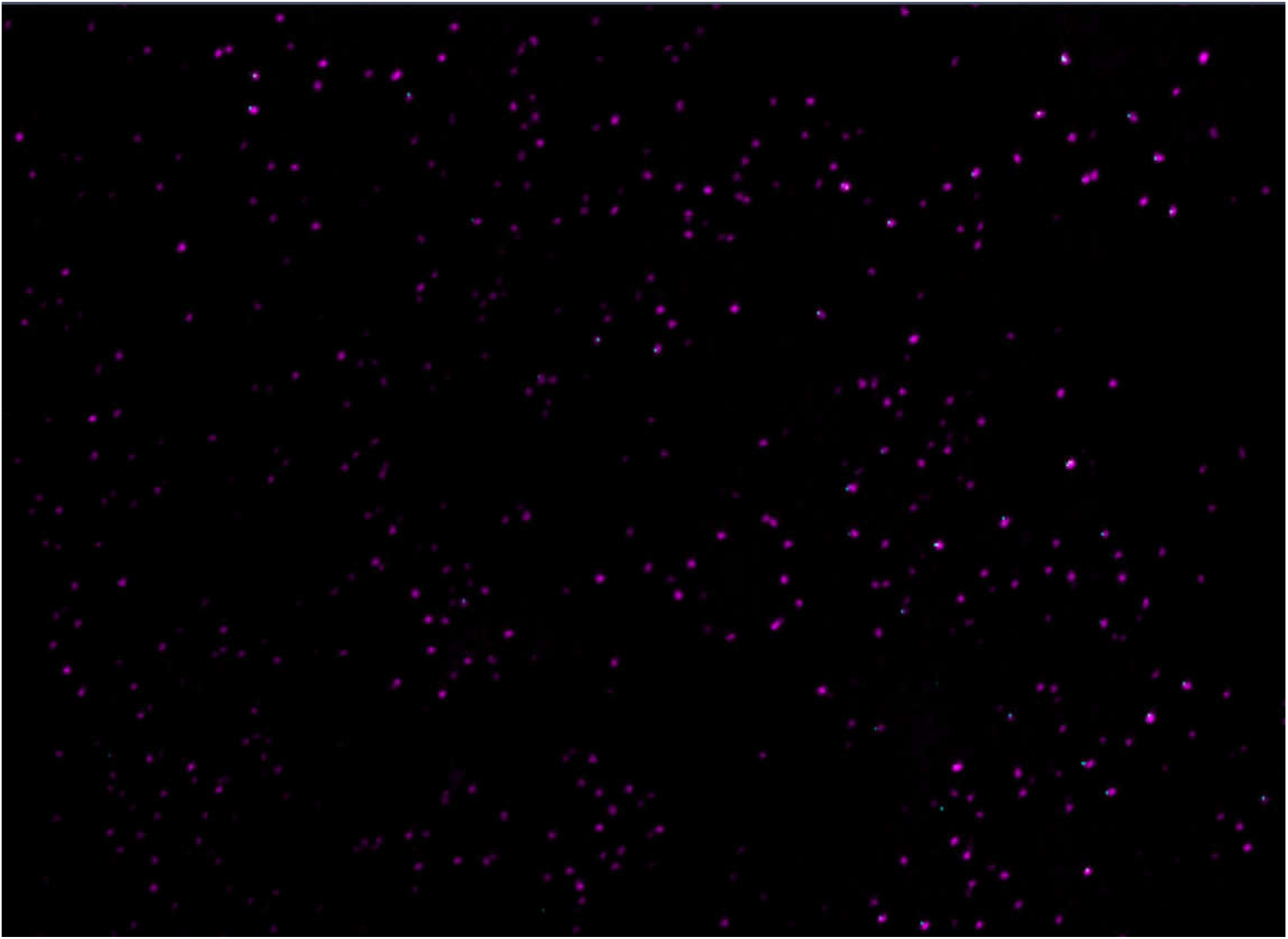
Co-staining of the lipid bilayer of urinary extracellular vesicles with MemGlow^TM^ 560 (purple) and the podocyte-specific marker podocalyxin with the antibody PODXL488 (cyan).

**Supplementary Figure 2.**
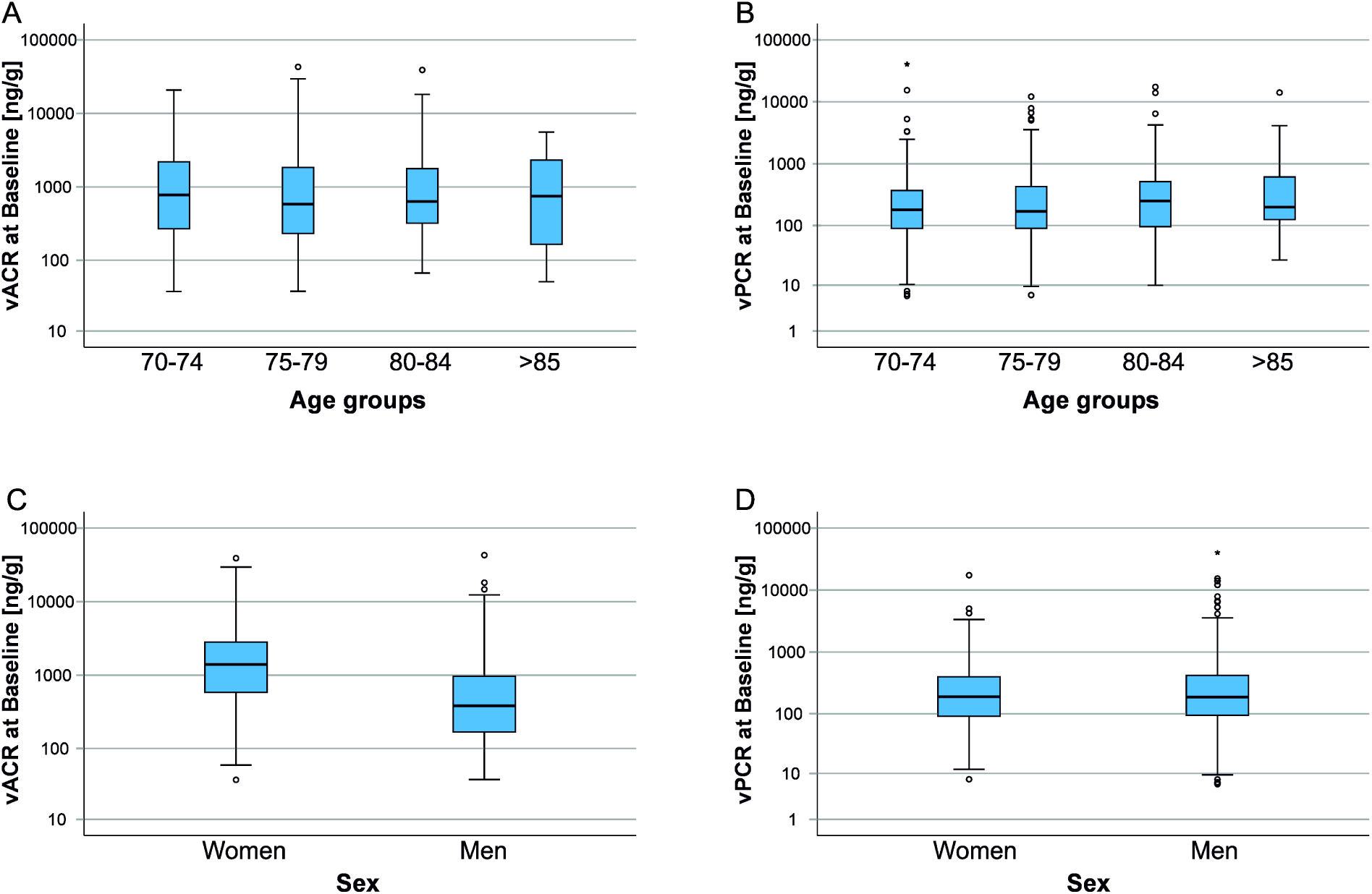
Box plots visualising vACR (left) and vPCR (right) distributions (log-scale on y axis) at baseline over age groups (A, B) and by sex (C, D). vPCR significantly differed by age and vACR by sex.

**Supplementary Figure 3.**
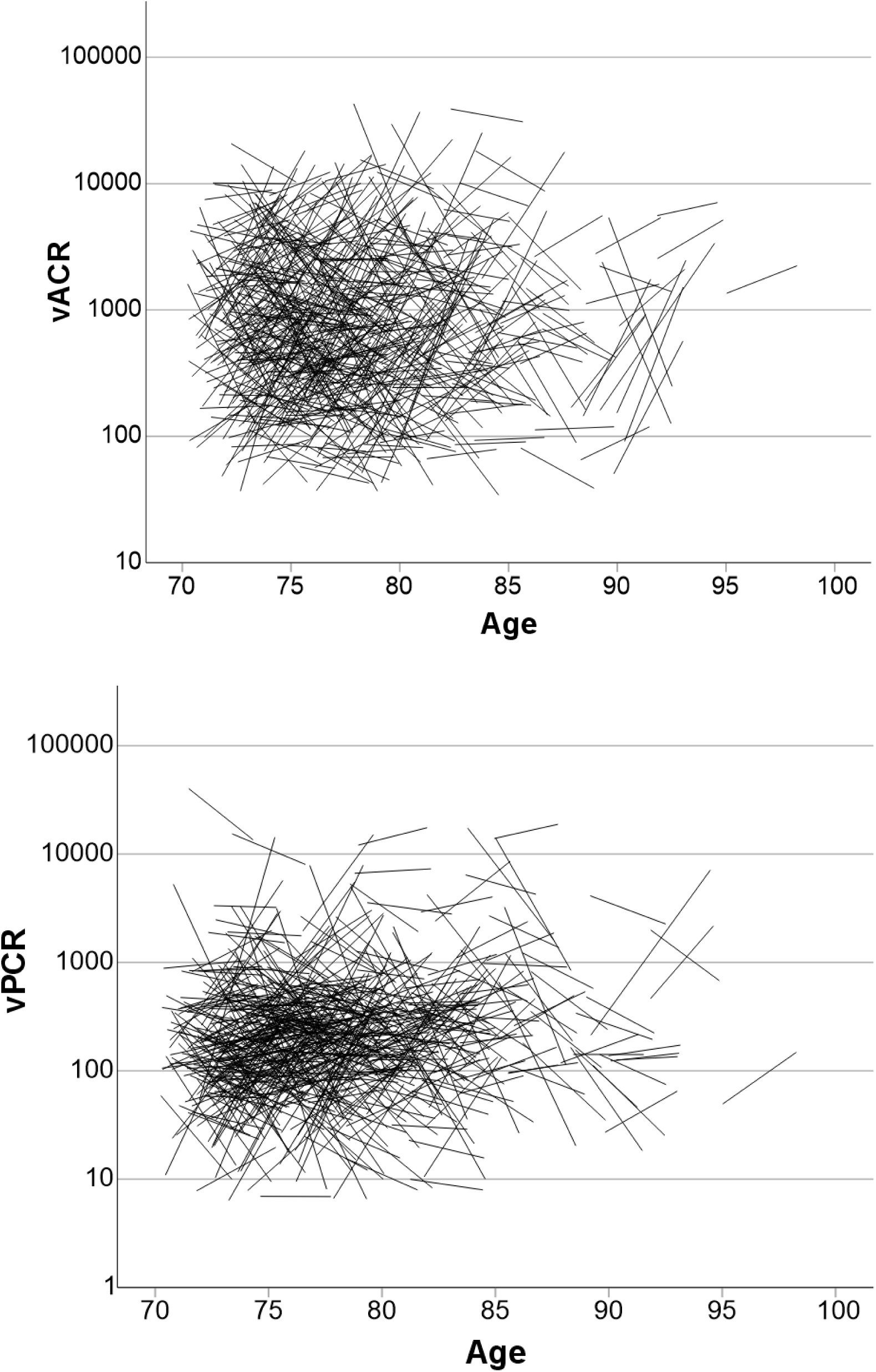
Spaghetti plots for change in vACR (top) and vPCR (bottom) between baseline and three years follow-up over baseline age.

**Supplementary Table 1.** Intra- and inter-assay CVs of triplicate measurements of vesicular albumin and podocalyxin.

|  | Vesicular albumin<br>(n = 26) | Vesicular podocalyxin<br>(n = 26) |
| --- | --- | --- |
| Intra-assay CV | 5.29% (0.06 – 16.86%)<br>9.76% (0.64 – 49.88%) | 5.53% (0.95 – 16.14%)<br>4.78% (1.60 – 13.85%) |
| Inter-assay CV (within one week) | 16.76% (0.51 – 137.62%) | 5.98% (0.57 – 23.29%) |
| Inter-assay CV (after 15 – 17 months) | 27.28% (8.57 – 166.77%) | 13.12% (2.96 – 37.13%) |

Mean (Min – Max) of individual CVs.

## Notes

### Competing Interest Statement

The authors have declared no competing interest.

### Author Declarations

Institutional Review Board of the University of Regensburg gave ethical approval for this work (IRB number: 12-101-0258).

## REFERENCES

1. Kidney Disease: Improving Global Outcomes (KDIGO) CKD Work Group (2024) KDIGO 2024 Clinical Practice Guideline for the Evaluation and Management of Chronic Kidney Disease. Kidney Int 105:S117–S314. 10.1016/j.kint.2023.10.018

2. Kovesdy CP (2022) Epidemiology of chronic kidney disease: an update 2022. Kidney Int Suppl (2011) 12:7–11. 10.1016/j.kisu.2021.11.003

3. Birn H, Christensen EI (2006) Renal albumin absorption in physiology and pathology. Kidney Int 69:440–449. 10.1038/sj.ki.5000141

4. Ballermann BJ, Nyström J, Haraldsson B (2021) The Glomerular Endothelium Restricts Albumin Filtration. Front Med (Lausanne) 8:766689. 10.3389/fmed.2021.766689

5. Zhuo JL, Li XC (2013) Proximal nephron. Compr Physiol 3:1079–1123. 10.1002/cphy.c110061

6. Dickson LE, Wagner MC, Sandoval RM et al. (2014) The proximal tubule and albuminuria: really! J Am Soc Nephrol 25:443–453. 10.1681/ASN.2013090950

7. van der Velde M, Matsushita K, Coresh J, et al. (2011) Lower estimated glomerular filtration rate and higher albuminuria are associated with all-cause and cardiovascular mortality. A collaborative meta-analysis of high-risk population cohorts. Kidney Int 79:1341–1352. 10.1038/ki.2010.536

8. Claudel SE, Verma A (2025) Albuminuria in Cardiovascular, Kidney, and Metabolic Disorders: A State-of-the-Art Review. Circulation 151:716–732. 10.1161/CIRCULATIONAHA.124.071079

9. Porrini E, Ruggenenti P, Luis-Lima S et al. (2019) Estimated GFR: time for a critical appraisal. Nat Rev Nephrol 15:177–190. 10.1038/s41581-018-0080-9

10. Erdbrügger U, Blijdorp CJ, Bijnsdorp IV et al. (2021) Urinary extracellular vesicles: A position paper by the Urine Task Force of the International Society for Extracellular Vesicles. J Extracell Vesicles 10:e12093. 10.1002/jev2.12093

11. Welsh JA, Goberdhan DCI, O’Driscoll L et al. (2024) Minimal information for studies of extracellular vesicles (MISEV2023): From basic to advanced approaches. J Extracell Vesicles 13:e12404. 10.1002/jev2.12404

12. Burger D, Thibodeau J-F, Holterman CE et al. (2014) Urinary podocyte microparticles identify prealbuminuric diabetic glomerular injury. J Am Soc Nephrol 25:1401–1407. 10.1681/ASN.2013070763

13. Cricrì G, Bellucci L, Montini G et al. (2021) Urinary Extracellular Vesicles: Uncovering the Basis of the Pathological Processes in Kidney-Related Diseases. Int J Mol Sci 22. 10.3390/ijms22126507

14. Schießl IM, Hammer A, Kattler V et al. (2016) Intravital Imaging Reveals Angiotensin II-Induced Transcytosis of Albumin by Podocytes. J Am Soc Nephrol 27:731–744. 10.1681/ASN.2014111125

15. Castrop H, Schießl IM (2017) Novel routes of albumin passage across the glomerular filtration barrier. Acta Physiol (Oxf) 219:544–553. 10.1111/apha.12760

16. Stark K, Olden M, Brandl C et al. (2015) The German AugUR study: study protocol of a prospective study to investigate chronic diseases in the elderly. BMC Geriatr 15:130. 10.1186/s12877-015-0122-0

17. Brandl C, Zimmermann ME, Günther F et al. (2018) On the impact of different approaches to classify age-related macular degeneration: Results from the German AugUR study. Sci Rep 8:8675. 10.1038/s41598-018-26629-5

18. Brandl C, Brücklmayer C, Günther F et al. (2019) Retinal Layer Thicknesses in Early Age-Related Macular Degeneration: Results From the German AugUR Study. Invest Ophthalmol Vis Sci 60:1581–1594. 10.1167/iovs.18-25332

19. Steinkirchner AB, Zimmermann ME, Donhauser FJ et al. (2022) Self-report of chronic diseases in old-aged individuals: extent of agreement with general practitioner medical records in the German AugUR study. J Epidemiol Community Health 76:931–938. 10.1136/jech-2022-219096

20. Donhauser FJ, Zimmermann ME, Steinkirchner AB et al. (2023) Cardiovascular Risk Factor Control in 70- to 95-Year-Old Individuals: Cross-Sectional Results from the Population-Based AugUR Study. J Clin Med 12. 10.3390/jcm12062102

21. Muli S, Meisinger C, Heier M et al. (2020) Prevalence, awareness, treatment, and control of hypertension in older people: results from the population-based KORA-age 1 study. BMC Public Health 20:1049. 10.1186/s12889-020-09165-8

22. Meisinger C, Döring A, Heier M et al. (2005) Type 2 diabetes mellitus in Augsburg--an epidemiological overview. Gesundheitswesen 67 Suppl 1:S103–9. 10.1055/s-2005-858251

23. Inker LA, Eneanya ND, Coresh J et al. (2021) New Creatinine- and Cystatin C-Based Equations to Estimate GFR without Race. N Engl J Med 385:1737–1749. 10.1056/NEJMoa2102953

24. Collot M, Ashokkumar P, Anton H et al. (2019) MemBright: A Family of Fluorescent Membrane Probes for Advanced Cellular Imaging and Neuroscience. Cell Chem Biol 26:600–614.e7. 10.1016/j.chembiol.2019.01.009

25. Webster AC, Nagler EV, Morton RL et al. (2017) Chronic Kidney Disease. Lancet 389:1238–1252. 10.1016/S0140-6736(16)32064-5

26. Levin A, Stevens PE (2011) Early detection of CKD: the benefits, limitations and effects on prognosis. Nat Rev Nephrol 7:446–457. 10.1038/nrneph.2011.86

27. Ardalan M, Hosseiniyan Khatibi SM, Rahbar Saadat Y et al. (2022) Migrasomes and exosomes; different types of messaging vesicles in podocytes. Cell Biol Int 46:52–62. 10.1002/cbin.11711

28. Lv L-L, Cao Y-H, Pan M-M et al. (2014) CD2AP mRNA in urinary exosome as biomarker of kidney disease. Clin Chim Acta 428:26–31. 10.1016/j.cca.2013.10.003

29. Yang R, Zhang H, Chen S et al. (2024) Quantification of urinary podocyte-derived migrasomes for the diagnosis of kidney disease. J Extracell Vesicles 13:e12460. 10.1002/jev2.12460

30. Lee H, Han KH, Lee SE et al. (2012) Urinary exosomal WT1 in childhood nephrotic syndrome. Pediatr Nephrol 27:317–320. 10.1007/s00467-011-2035-2

31. Kalani A, Mohan A, Godbole MM et al. (2013) Wilm’s tumor-1 protein levels in urinary exosomes from diabetic patients with or without proteinuria. PLoS One 8:e60177. 10.1371/journal.pone.0060177

32. Lu J, Hu ZB, Chen PP et al. (2019) Urinary podocyte microparticles are associated with disease activity and renal injury in systemic lupus erythematosus. BMC nephrology 20:1–9. 10.1186/s12882-019-1482-z

33. Kwon SH, Woollard JR, Saad A et al. (2017) Elevated urinary podocyte-derived extracellular microvesicles in renovascular hypertensive patients. Nephrol Dial Transplant 32:800–807. 10.1093/ndt/gfw077

34. Abe H, Sakurai A, Ono H et al. (2018) Urinary Exosomal mRNA of WT1 as Diagnostic and Prognostic Biomarker for Diabetic Nephropathy. J Med Invest 65:208–215. 10.2152/jmi.65.208

35. Zhou H, Kajiyama H, Tsuji T et al. (2013) Urinary exosomal Wilms’ tumor-1 as a potential biomarker for podocyte injury. Am J Physiol Renal Physiol 305:F553–9. 10.1152/ajprenal.00056.2013

36. Miller D, Eagle-Hemming B, Sheikh S et al. (2022) Urinary extracellular vesicles and micro-RNA as markers of acute kidney injury after cardiac surgery. Sci Rep 12:10402. 10.1038/s41598-022-13849-z

37. Hara M, Yamagata K, Tomino Y et al. (2012) Urinary podocalyxin is an early marker for podocyte injury in patients with diabetes: establishment of a highly sensitive ELISA to detect urinary podocalyxin. Diabetologia 55:2913–2919. 10.1007/s00125-012-2661-7

38. He F-F, Gong Y, Li Z-Q et al. (2018) A New Pathogenesis of Albuminuria: Role of Transcytosis. Cell Physiol Biochem 47:1274–1286. 10.1159/000490223

39. Akilesh S, Huber TB, Wu H et al. (2008) Podocytes use FcRn to clear IgG from the glomerular basement membrane. Proc Natl Acad Sci U S A 105:967–972. 10.1073/pnas.0711515105

40. Lu J, Hu Z-B, Chen P-P et al. (2019) Urinary levels of podocyte-derived microparticles are associated with the progression of chronic kidney disease. Ann Transl Med 7:445. 10.21037/atm.2019.08.78

41. Okamura K, Dummer P, Kopp J et al. (2013) Endocytosis of albumin by podocytes elicits an inflammatory response and induces apoptotic cell death. PLoS One 8:e54817. 10.1371/journal.pone.0054817

42. Yoshida S, Nagase M, Shibata S et al. (2008) Podocyte injury induced by albumin overload in vivo and in vitro: involvement of TGF-beta and p38 MAPK. Nephron Exp Nephrol 108:e57–68. 10.1159/000124236

43. Pawluczyk IZA, Pervez A, Ghaderi Najafabadi M et al. (2014) The effect of albumin on podocytes: the role of the fatty acid moiety and the potential role of CD36 scavenger receptor. Exp Cell Res 326:251–258. 10.1016/j.yexcr.2014.04.016

44. Chen S, He F-F, Wang H et al. (2011) Calcium entry via TRPC6 mediates albumin overload-induced endoplasmic reticulum stress and apoptosis in podocytes. Cell Calcium 50:523–529. 10.1016/j.ceca.2011.08.008

45. He F-F, Zhang C, Chen S et al. (2011) Role of CD2-associated protein in albumin overload-induced apoptosis in podocytes. Cell Biol Int 35:827–834. 10.1042/CBI20100411

46. Blaine J, Dylewski J (2020) Regulation of the Actin Cytoskeleton in Podocytes. Cells 9. 10.3390/cells9071700

47. Singh P, Szigyártó IC, Ricci M et al. (2023) Removal and identification of external protein corona members from RBC-derived extracellular vesicles by surface manipulating antimicrobial peptides. J Extracell Biol 2:e78. 10.1002/jex2.78

48. Dietz L, Oberländer J, Mateos-Maroto A et al. (2023) Uptake of extracellular vesicles into immune cells is enhanced by the protein corona. J Extracell Vesicles 12:e12399. 10.1002/jev2.12399

49. Liam-Or R, Faruqu FN, Walters A et al. (2024) Cellular uptake and in vivo distribution of mesenchymal-stem-cell-derived extracellular vesicles are protein corona dependent. Nature nanotechnology:1–10. 10.1038/s41565-023-01585-y

50. Wolf M, Poupardin RW, Ebner-Peking P et al. (2022) A functional corona around extracellular vesicles enhances angiogenesis, skin regeneration and immunomodulation. J Extracell Vesicles 11:e12207. 10.1002/jev2.12207

51. Gomes FG, Andrade AC, Wolf M et al. (2022) Synergy of Human Platelet-Derived Extracellular Vesicles with Secretome Proteins Promotes Regenerative Functions. Biomedicines 10. 10.3390/biomedicines10020238

52. Tóth EÁ, Turiák L, Visnovitz T et al. (2021) Formation of a protein corona on the surface of extracellular vesicles in blood plasma. J Extracell Vesicles 10:e12140. 10.1002/jev2.12140

53. Liangsupree T, Multia E, Riekkola M-L (2021) Modern isolation and separation techniques for extracellular vesicles. J Chromatogr A 1636:461773. 10.1016/j.chroma.2020.461773

54. Maggio S, Polidori E, Ceccaroli P et al. (2021) Current Methods for the Isolation of Urinary Extracellular Vesicles. Methods Mol Biol 2292:153–172. 10.1007/978-1-0716-1354-2_14

55. Hogan MC, Johnson KL, Zenka RM et al. (2014) Subfractionation, characterization, and in-depth proteomic analysis of glomerular membrane vesicles in human urine. Kidney Int 85:1225–1237. 10.1038/ki.2013.422

